# Infection and mortality of healthcare workers worldwide from COVID-19: a scoping review

**DOI:** 10.1101/2020.06.04.20119594

**Authors:** Soham Bandyopadhyay, Ronnie E Baticulon, Murtaza Kadhum, Muath Alser, Daniel K Ojuka, Yara Badereddin, Archith Kamath, Sai Arathi Parepalli, Grace Brown, Sara Iharchane, Sofia Gandino, Zara Markovic-Obiago, Samuel Scott, Emery Manirambona, Asif Machhada, Aditi Aggarwal, Lydia Benazaize, Mina Ibrahim, David Kim, Isabel Tol, Elliott H Taylor, Alexandra Knighton, Dorothy Bbaale, Duha Jasim, Heba Alghoul, Henna Reddy, Hibatullah Abuelgasim, Alicia Sigler, Kirandeep Saini, Leenah Abuelgasim, Mario Moran-Romero, Mary Kumarendran, Najlaa Abu Jamie, Omaima Ali, Raghav Sudarshan, Riley Dean, Rumi Kisyova, Sonam Kelzang, Sophie Roche, Tazin Ahsan, Yethrib Mohamed, Andile Maqhawe Dube, Grace Paidamoyo Gwini, Rashidah Gwokyalya, Robin Brown, Mohammad Rabiul Karim Khan Papon, Zoe Li, Salvador Sun Ruzats, Somy Charuvila, Noel Peter, Khalil Khalidy, Nkosikhona Moyo, Osaid Alser, Arielis Solano, Eduardo Robles-Perez, Aiman Tariq, Mariam Gaddah, Spyros Kolovos, Faith C Muchemwa, Abdullah Saleh, Amanda Gosman, Rafael Pinedo-Villanueva, Anant Jani, Roba Khundkar

## Abstract

**Objectives:** To estimate COVID-19 infections and deaths in healthcare workers (HCWs) from a global perspective.

**Design:** Scoping review.

**Methods:** Two parallel searches of academic bibliographic databases and grey literature were undertaken. Governments were also contacted for further information where possible. Due to the time-sensitive nature of the review and the need to report the most up-to-date information for an ever-evolving situation, there were no restrictions on language, information sources utilised, publication status, and types of sources of evidence. The AACODS checklist was used to appraise each source of evidence.

**Outcome measures:** Publication characteristics, country-specific data points, COVID-19 specific data, demographics of affected HCWs, and public health measures employed

**Results:** A total of 152,888 infections and 1413 deaths were reported. Infections were mainly in women (71.6%) and nurses (38.6%), but deaths were mainly in men (70.8%) and doctors (51.4%). Limited data suggested that general practitioners and mental health nurses were the highest risk specialities for deaths. There were 37.17 deaths reported per 100 infections for healthcare workers aged over 70. Europe had the highest absolute numbers of reported infections (119628) and deaths (712), but the Eastern Mediterranean region had the highest number of reported deaths per 100 infections (5.7).

**Conclusions:** HCW COVID-19 infections and deaths follow that of the general world population. The reasons for gender and speciality differences require further exploration, as do the low rates reported from Africa and India. Although physicians working in certain specialities may be considered high-risk due to exposure to oronasal secretions, the risk to other specialities must not be underestimated. Elderly HCWs may require assigning to less risky settings such as telemedicine, or administrative positions. Our pragmatic approach provides general trends, and highlights the need for universal guidelines for testing and reporting of infections in HCWs.

**Summary Box:** *What is already known on this topic:* In China, studies documented over 3,300 confirmed cases of infected healthcare workers in early March. In the United States, as high as 19% of patients had been identified as healthcare workers. There are no studies that perform a global examination of COVID-19 infections and deaths in the health workforce.

*What this study adds:* To our knowledge, this is the first study assessing the number of healthcare workers who have been infected with or died from COVID-19 globally. The data from our study suggest that although infections were mainly in women and nurses, COVID-19 related deaths were mainly in men and doctors; in addition, our study found that Europe had the highest numbers of infection and death, but the lowest case-fatality-rate, while the Eastern Mediterranean had the highest case-fatality-rate.

## 1. Introduction

From a cluster of patients with pneumonia linked to a wet market in Wuhan, China in late December 2019, the coronavirus disease (COVID-19) has rapidly evolved into a full-blown pandemic (1,2). At the time of writing (23^rd^ May), over five million people have been infected across 213 countries and territories, leading to more than 300,000 deaths worldwide (3). On the frontlines of this global crisis are healthcare workers (HCWs) with the substantial task of diagnosing and treating an exponentially growing number of acutely ill patients, often having to make critical decisions under physical and psychological pressure (4–6). The World Health Organization (WHO) defines health workers as “all people engaged in actions whose primary intent is to enhance health” (7). This encompasses doctors, nurses, midwives, paramedical staff, hospital administrators and support staff, and community workers, all of whom now face the occupational risk of becoming infected with COVID-19, and at worst, even death.

In one of the first cohorts of COVID-19 patients from Wuhan, 40/138 (29%) were medical staff (8). A later study documented 1,716 infected HCWs among 44,672 confirmed cases in China, with the number rising to over 3,300 in early March (9,10). In the United States, as high as 19% of patients had been identified as HCWs (11). These figures are reminiscent of the 2002 to 2004 severe acute respiratory syndrome (SARS) outbreak, in which 11 to 57% of total cases in affected countries were HCWs, equivalent to one in five patients overall (12). Onward transmission of SARS occurred mainly through HCWs and within the healthcare setting, emphasizing the need for appropriate personal protective equipment (PPE) and adherence to infection control principles in the containment and eradication of emerging diseases (12–16).

Ensuring the protection of HCWs is a crucial element of any country’s strategic response to the COVID-19 crisis, especially as governments rush to increase healthcare capacity to cope with the surge of patients requiring urgent care. The WHO has issued recommendations on the rational use of PPE in hospital and community settings (17). Several colleges and speciality societies have formulated algorithms and guidelines to decrease the risk of COVID-19 transmission in their fields of practice (18–22). Nevertheless, protecting HCWs remains a challenge for most countries, where shortages of adequate PPE is a daily concern. Limited testing capacity precludes early identification and isolation of cases, leading to unnecessary occupational exposure for HCWs, particularly since a high number of COVID-19 patients remain asymptomatic. In a vicious cycle, shortages of HCWs may compel staff to continue working for days on end, even under fatigue or when symptoms manifest, further increasing the risk of transmission.

Unmitigated, rising infection and mortality rates in HCWs will not only paralyse a country’s response to COVID-19. It is bound to have significant, long-term impact in healthcare delivery, particularly in health systems already grappling with workforce shortage due to lack of trained personnel, skilled labour migration, and geographical maldistribution, even prior to pandemic times (23–26).

There are no studies that examine COVID-19 infection and death amongst HCWs globally. The aim of this scoping review was to, therefore, estimate the number of COVID-19 infections and deaths in HCWs in every country in the world, with further demographical analyses where data was available.

## 2. Methods

### 2.1 Overview

A scoping review on the number and proportion of HCWs who have been infected with or died from COVID-19 was conducted as per the published and registered protocol (27) (Appendix S1).

The primary outcomes of interest were COVID-19 infections and deaths in HCWs worldwide. Subgroup analyses were performed according to WHO region, country, and demographic characteristics.

Frameworks published by Arksey and O’Malley (28), Levac et al. (29), and Joana Briggs Institute (30) were used to guide the scoping review. The Preferred Reporting Items for Systemic Review and Meta-Analyses extension for Scoping Reviews (PRISMA-ScR) guidelines (31) were used to report the findings (Appendix S2).

Due to the time-sensitive nature of the review and the need to report the most up-to-date information for an ever-evolving situation, there were no restrictions on language, information sources utilised, publication status, and types of sources of evidence. Prior to the commencement of this study, all reviewers attended an online training and support session to ensure an accurate and standardised approach to the overall methodological process. Ongoing research support was provided for all collaborators throughout the process.

### 2.2 Search Strategy

Two parallel searches of academic bibliographic databases and grey literature were undertaken.

1. Bibliographic search: The search protocol for this scoping review was executed in MEDLINE and EMBASE, covering the period between the first reported case in the world on 17^th^ November 2019 to 8^th^ May 2020. The search strategy used variants and combinations of search terms related to HCWs and COVID-19 (Appendix S3). The retrieved studies were exported, and duplicate articles were discarded. Two reviewers independently screened the titles and abstracts based on the pre-defined inclusion and exclusion criteria (Appendix S1). The full texts of the remaining articles were retrieved and screened by two reviewers independently. Any disagreements were resolved by a third reviewer. The reference lists of all included articles were scrutinised to locate additional relevant publications not identified during the database searches. The reviewers also consulted with senior HCWs – identified and approached through the network of the Oxford University Global Surgery Group – across the world to identify additional publications.
2. Grey literature search: A grey literature search was performed to include sources dedicated to COVID-19 or world data. These sources included government websites, non-governmental websites, social media sites, media websites, and preprints on medRxiv (32). Snowball searching using a web-based search engine (Google) was utilised to find additional documents and online sources. A pragmatic approach was taken for the grey literature search and a stepwise guide was provided to all data collectors to ensure consistency of search strategy. A full record of the conducted search is provided in Appendix S4. The reference list of all included documents identified through the grey literature search was examined to identify any further relevant documents and online sources missed through the above search strategy, until a saturation point was reached where no new sources were identified.

All searches were completed in duplicate by two reviewers independently. A third reviewer validated these searches and resolved any disparities when they arose. Lastly, governments were contacted for further information where possible. The initial search was completed for 22/04/2020. The search was then updated for 08/05/2020 for primary and secondary outcomes.

### 2.3 Data extraction

A data extraction form (Appendix S5) was developed to collect the information necessary for data synthesis. This form was piloted by the team before its use. Data extraction was completed in duplicate by two reviewers independently. A third reviewer validated the data extracted and resolved any disagreements. Several data points were extracted, including publication characteristics, country-specific data points, COVID-19 specific data, demographics of affected HCWs and public health measures employed (Table 1).

**Table 1:** The type and specific data points extracted in the scoping review process.

| **Data extracted** |  |
| --- | --- |
| **Publication characteristics** | Title of publication<br>Year published<br>Authors<br>Country of origin<br>Type of publication |
| **Country-specific data** | Total number of healthcare workers<br>Total population<br>Number of total hospital beds available<br>Number of intensive care unit beds and ventilators available |
| **COVID-19 specific data** | Number of nationwide cases<br>Number of COVID-19 tests performed<br>Number of intensive care unit admissions<br>Mortality rates<br>Average length time for symptoms and admission |
| **Demographics of affected healthcare workers** | Age, sex, ethnicity, stage of training and role of each affected healthcare worker |
| **Public health measures** | Social distancing strategies<br>Testing, self-isolation and contact tracing<br>Travel restrictions<br>Stay at home orders |
| **Publication characteristics** | Title of publication<br>Year published<br>Authors<br>Country of origin<br>Type of publication |

### 2.4 Quality assessment

Due to the heterogeneity of the various sources of evidence used in this scoping review, each source of evidence was critically appraised. Two reviewers independently classified the quality of each included document using the AACODS checklist (Appendix S6). Any uncertainty regarding the quality of an included document was resolved through discussion among the reviewers.

### 2.5 Synthesis of results

Mortality and infection numbers have been pooled. HCW infection and deaths due to COVID-19 as a proportion of total population infections and deaths respectively due to COVID-19 have been calculated. HCW deaths due to COVID-19 as a proportion of all HCW COVID-19 infections have also been calculated and expressed as a case fatality rate (CFR): number of reported deaths per 100 cases of reported infections.

### 2.5 Patient and Public Involvement

Patient and public involvement was not appropriate for this scoping review.

## 3. Results

### 3.1 Search & Selection of Studies/sources

Searches were conducted up to 08/05/2020. The searches yielded a total of 976 potentially relevant citations. After data validation and cleansing, duplicated and irrelevant citations were removed manually. At this point, 594 citations met the eligibility criteria based on our protocol (See PRISMA Diagram Figure 1).

**Fig 1.**
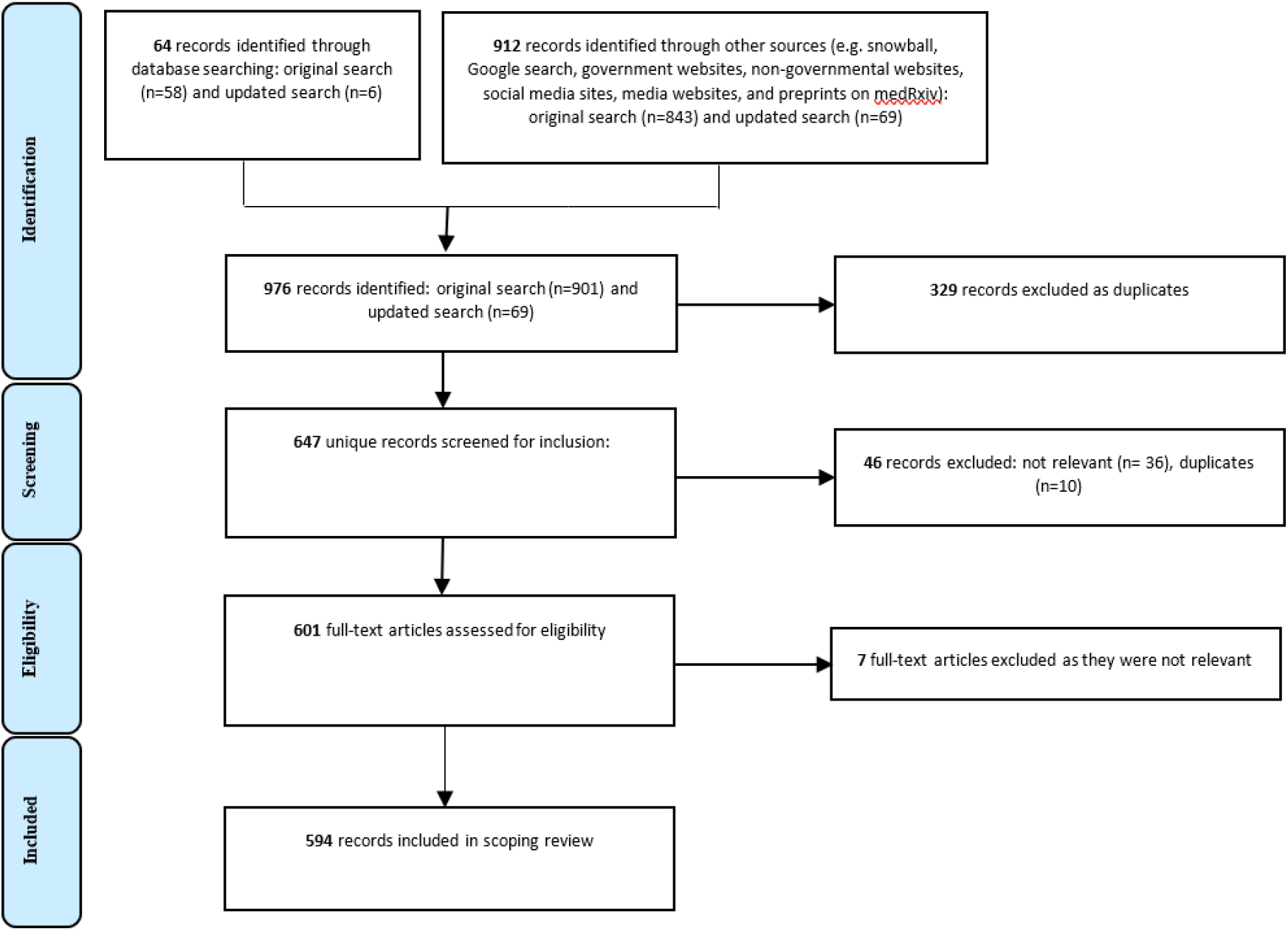
PRISMA flowchart of the source selection process.

More specifically for the bibliographic database search a total of 64 articles were retrieved. Ten duplicates were detected, thus 54 studies remained. After the screening of titles and abstracts based on inclusion and exclusion criteria, 15 of these progressed to full-text screening and eight studies were included in the final analysis. The grey literature search yielded 912 citations. After screening for inclusion and eligibility, 586 citations remained and were included in the study.

### 3.2 Characteristics of included sources

Characteristics of the included sources are described in Table 2. Overall, 594 records were included, of which 14 were journal articles. Of the remaining records, 19.5% (n=116) were government documents, 16.5% (n=98) were government websites, 48.3% (n=287) were media articles, 9.6% (n=57) were research or academic reports, 3% (n=18) were statistical websites, and 0.7% (n=4) were primary sources. All sources were appraised per the ACCODS checklist, as shown in appendix S4. Primary outcomes were available for 85.1% of all countries (166/195).

**Table 2.** Characteristics of included sources.

|  |  | Number | Percentage (%) |
| --- | --- | --- | --- |
| **Publication type** | Journal Article | 14 | 2.4 |
|  | Government Document | 116 | 19.5 |
|  | Government Websites | 98 | 16.5 |
|  | Media Articles | 287 | 48.3 |
|  | Research/Academic Reports | 57 | 9.6 |
|  | Statistical Websites | 18 | 3.0 |
|  | Primary Sources | 4 | 0.7 |
| **Countries with no data for primary outcome** | Angola | 29 | 14.9 |
|  | Barbados |  |  |
|  | Belize |  |  |
|  | Bolivia |  |  |
|  | Cote d'Ivoire |  |  |
|  | Djibouti |  |  |
|  | Dominica |  |  |
|  | Eritrea |  |  |
|  | Iraq |  |  |
|  | Jordan |  |  |
|  | Liechtenstein |  |  |
|  | Luxembourg |  |  |
|  | Malawi |  |  |
|  | Maldives |  |  |
|  | Mauritania |  |  |
|  | Monaco |  |  |
|  | Nicaragua |  |  |
|  | North Korea |  |  |
|  | North Macedonia (formerly Macedonia) |  |  |
|  | Norway |  |  |
|  | Oman |  |  |
|  | Paraguay |  |  |
|  | Qatar |  |  |
|  | Slovakia |  |  |
|  | Solomon Islands |  |  |
|  | Syria |  |  |
|  | Tonga |  |  |
|  | Turkmenistan |  |  |
|  | United Arab Emirates (UAE) |  |  |

### 3.3 Outcomes

#### 3.3.1 Primary outcomes

##### 3.3.1 a. Number of healthcare workers infected with COVID-19 worldwide

As of 08/05/2020, a total of 152,888 HCWs had been reported to have been infected with COVID-19 (Figure 1). This was 3.9% of the total number of 3,912,156 patients with COVID-19 worldwide (table 6). A total of 130 countries reported at least one case of HCW infection with COVID-19 (Figure 1 and Appendix S7).

**Fig 1:**
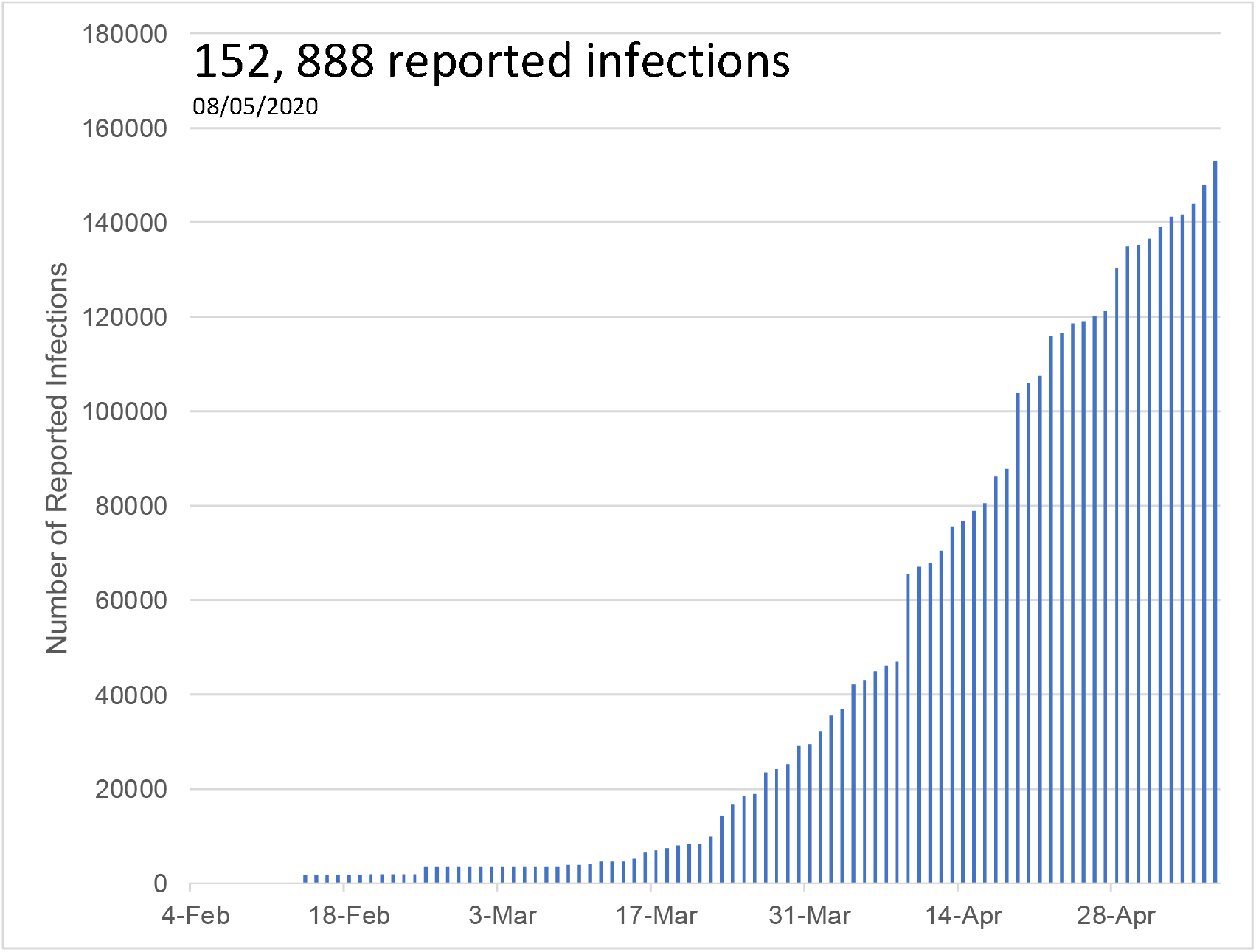
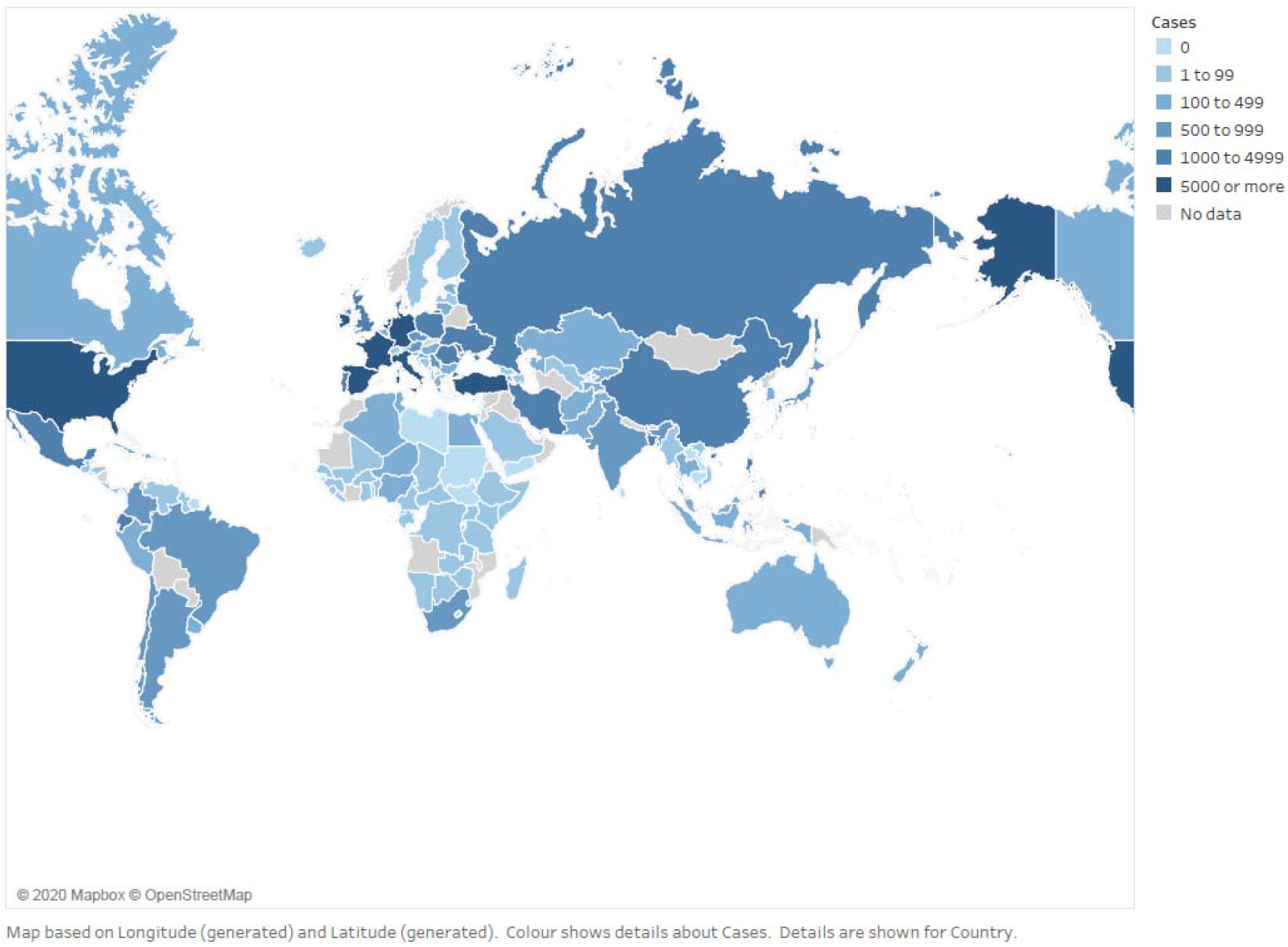
Cumulative number of reported COVID-19 infections in healthcare workers worldwide over time & total number of reported cases of COVID-19 infections in healthcare workers worldwide on 08/05/2020.

##### 3.3.1 b. Number of healthcare worker deaths with COVID-19 worldwide

The total number of reported HCW deaths as of 8th May 2020 was 1413 (Figure 2). This was 0.5% of the total number of 270, 426 COVID-19 deaths worldwide. This also suggests that for every hundred HCWs that got infected, one died. As of 8th May 2020, 67 countries had reported HCW deaths related to COVID-19 (Figure 2 and Appendix S7). China and Italy were the first two countries to report HCW deaths, and the first deaths in each of these countries occurred over a month apart (Appendix S8).

**Fig 2.**
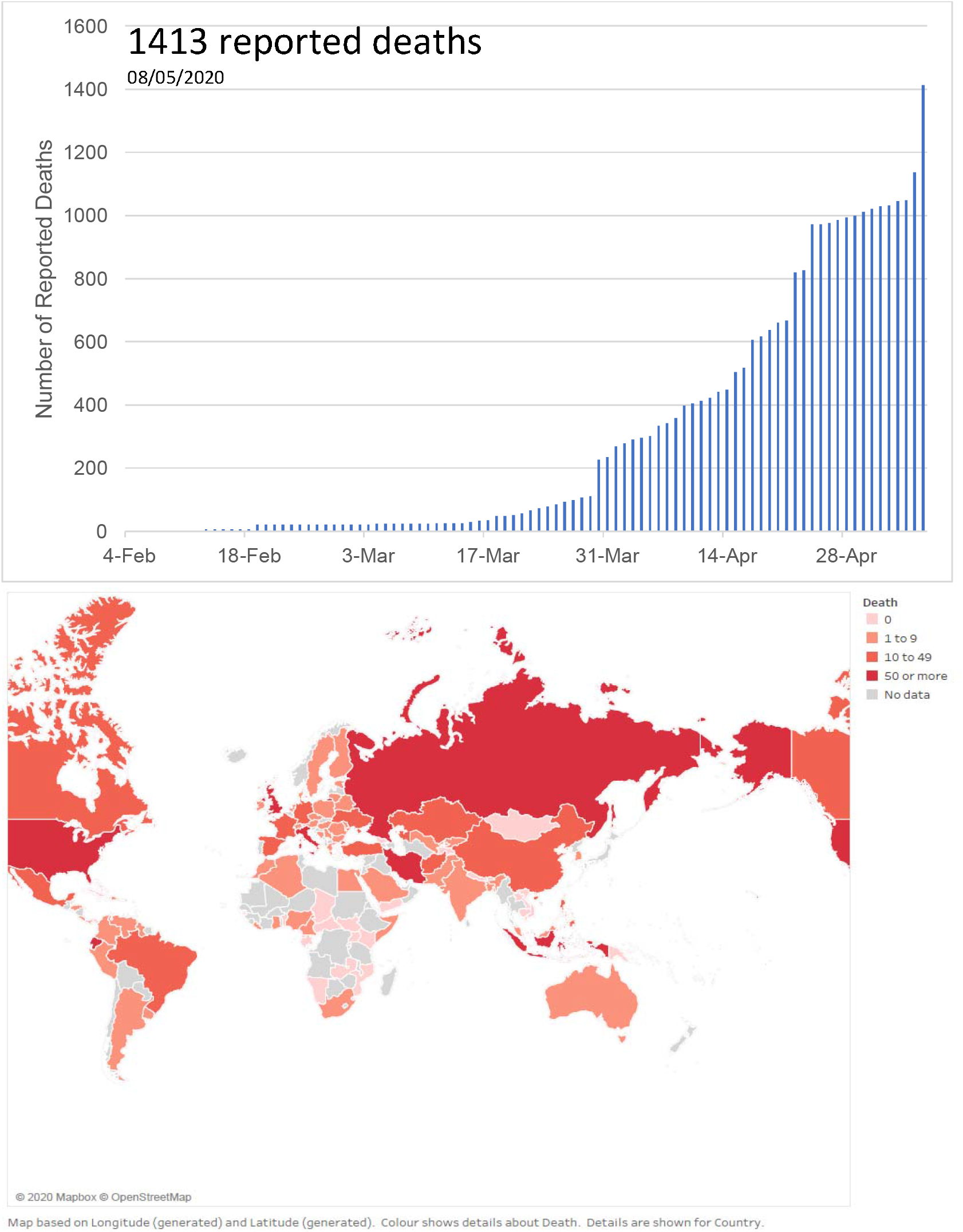
Cumulative number of reported COVID-19 deaths in healthcare workers worldwide & total number of reported cases of COVID-19 deaths in healthcare workers worldwide.

#### 3.3.2 Subgroup analysis

##### 3.3.2. a. Characteristics of healthcare workers who were infected with COVID-19

Although most countries released the total number of HCW deaths or infections, most published reports did not include details on the age category, ethnicity, or role of workers. The data below is based on a smaller number of countries that made this data available.

The overall median age of the reported HCWs who were infected was 47.3 years (range: 18-84), 71.6% of whom were women. The overall median age of the HCWs who died was 56.2 years (range: 18-84), 29.2% of them women. The CFR for men and women were 9.47 and 1.55, respectively. Nurses were the largest HCW group with COVID-19 infection (38.6% of those infected). However, doctors were the largest group of HCWs who died (51.4%). Ethnicity data for deaths were available for Australia, France, and the United Kingdom (UK) and showed 73 deaths in white HCWs and 106 deaths in non-white HCWs. Ethnicity data for infections were only available from the United States of America (USA) and showed 2743 infections in white HCWs and 1058 in non-white HCWs.

**Table 3:** Characteristics of healthcare workers with COVID-19 infection and deaths.

| Characteristic | Infection | Death |
| --- | --- | --- |
| **Age**<sup>i</sup> | Median Age: 47.3 yrs (n=14 058)<br>Range: 18 - 84 yrs | Median Age: 56.2 yrs (n = 343)<br>Range: 18 - 84 yrs |
| **Sex**<sup>ii</sup> | Male: 28.4% (n = 5806)<br>Female: 71.6% (n = 14656) | Male: 70.8% (n = 550)<br>Female: 29.2% (n = 227) |
| **Level (number)**<sup>iii</sup> | Student: <0.1% (n = 13)<br>Trainee: <0.1% (n = 2)<br>Qualified: 99.9% (n = 36081)<br>Retired: <0.1% (n = 5) | Student: 0.3% (n = 1)<br>Trainee: 0.6% (n = 2)<br>Qualified: 96.4% (n = 350)<br>Retired: 2.7% (n = 10) |
| **Type of HCW**<sup>iv</sup> | Doctor: 31.3% (n = 8688)<br>Nurse: 38.6% (n = 10706)<br>Midwives: <0.1% (n = 9)<br>Allied Health Professionals: 23.1% (n = 6394)<br>Support Staff: 6.8% (n = 1899)<br>Administrators: <0.1% (n = 27) | Doctor: 51.4% (n = 525)<br>Nurse: 25.3% (n = 259)<br>Midwives: 0.9% (n = 9)<br>Allied Health Professionals: 12.2% (n = 125)<br>Support Staff: 7.2% (n = 74)<br>Administrators: 2.8% (n = 29) |
| **Admission to intensive care unit**<sup>v</sup> | n = 1158 | NA |
|  | *i. Data from 15% of all countries; ii. Data from 21% of all countries; iii. Data from 7% of all countries; iv. Data from 13% of all countries; v. Data from 10% of all countries* | *i. Data from 17% of all countries; ii. Data from 21% of all countries; iii. Data from 13% of all countries; iv. Data from 22% of all countries;* |

##### 3.3.2. b Age

Age-stratified figures were not available for most countries. Data were only available for 14058 of the 152888 (9.2%) reported HCW infection cases and 343 of the 1413 (24.3%) reported HCW deaths as a result of COVID-19. Of all countries, 15% reported age-related information for COVID-19 cases and 17% reported information on COVID-19 deaths. The majority of infections was reported in the 50-59 year age range. The lowest reported number of infections were in the group aged over 70. However this age group had the highest CFR (Table 4).The majority of deaths were also reported in the 50-59 age range, with the lowest number reported in the 18-29 age group.

**Table 4:**
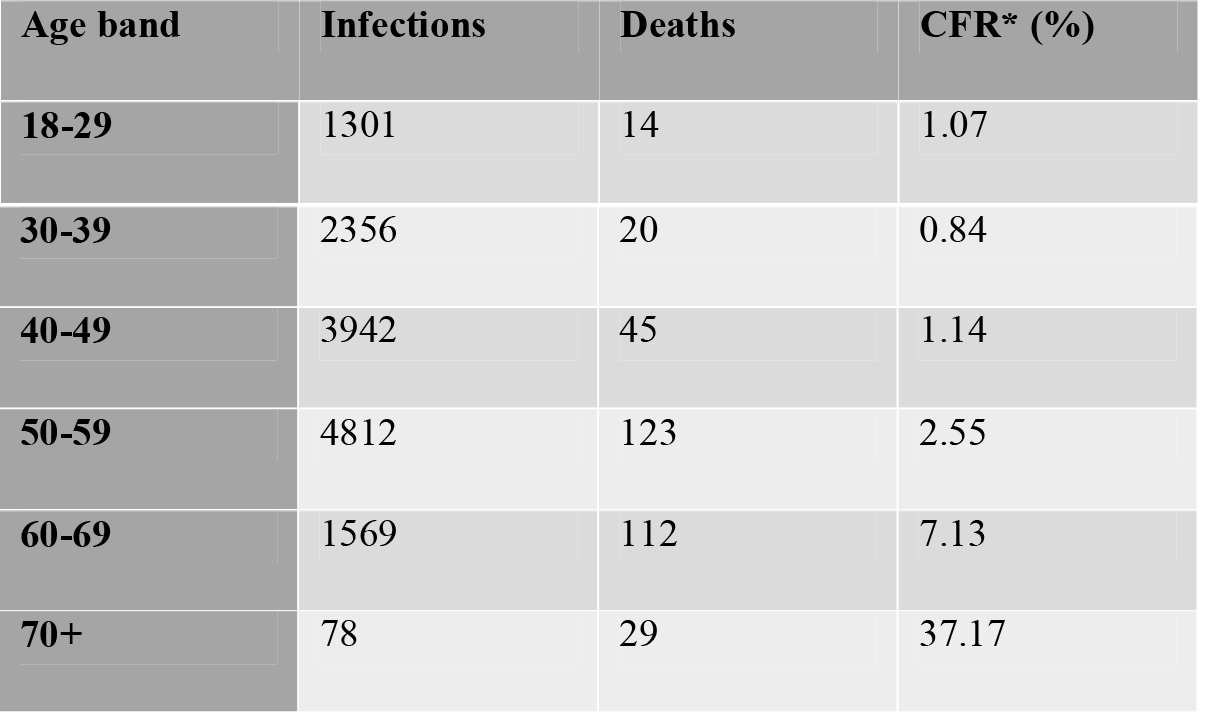
Distribution of infections, deaths and case-fatality in healthcare workers– data does not include cases with unknown age. *Case fatality rate is the number of reported deaths per 100 cases of reported infections.

| Age band | Infections | Deaths | CFR* (%) |
| --- | --- | --- | --- |
| **18-29** | 1301 | 14 | 1.07 |
| **30-39** | 2356 | 20 | 0.84 |
| **40-49** | 3942 | 45 | 1.14 |
| **50-59** | 4812 | 123 | 2.55 |
| **60-69** | 1569 | 112 | 7.13 |
| **70+** | 78 | 29 | 37.17 |

##### 3.3.2. c Specialities

Speciality-related data were only available from 14 countries: Argentina, Australia, Austria, Bahamas, Cameroon, Canada, China, Colombia, France, Germany, Ghana, Guyana, Turkey, and UK (13% of all data). General practitioners (GPs) appear to be the largest group of doctors who died while mental health nurses constituted the largest group of nurse subspecialists who lost their lives to COVID-19 (Table 5). There were 30 reported deaths in the UK amongst doctors, one third of which were GPs.

**Table 5:** Healthcare worker mortality by subspecialty (data available for only 13% of all deaths)

|  |  |  |
| --- | --- | --- |
| **Nurses** | Medicine: 15 | COVID-19 ward 4<br>Intensive care units 3<br>Acute admission 3<br>Adult care 2<br>Palliative 2<br>Cardiology 1 |
|  | Surgery: 1 | Orthopaedics 1 |
|  | Other: 26 | Mental health 14<br>Care home 8<br>Community 2<br>Dental 1<br>Disability 1 |
| **Doctors** | Medicine: 45 | General practitioner 18<br>Emergency medicine 5<br>Internal medicine 4<br>Paediatrics 3<br>Geriatrics 3<br>Neurology 2<br>Pathologist 2<br>Haem-oncology 1<br>Infectious disease 1<br>Microbiology 1<br>Nephrology 1<br>Psychiatry 1<br>Respiratory 1<br>Anaesthetics 1<br>Gastroenterology 1 |
|  | Surgery: 14 | Obstetrics and Gynaecology 3<br>Ophthalmology 2<br>Ear, Nose, and Throat 2<br>Urology 2<br>Cardiothoracic 1<br>Endocrine surgery 1<br>General surgery 1<br>Orthopaedics 1<br>Vascular 1 |
|  | Other: 6 | Director 2<br>Public health 1<br>Unknown 3 |

##### 3.3.2. d WHO Regions

The highest number of COVID-19 infections of HCWs were reported in Europe (119628, 78.2%), whilst the lowest number was reported in Africa (1472, 1.0%) (Figure 3). The same regional pattern was observed regarding deaths: Europe reported the highest number of deaths (712, 50.4%) and Africa the lowest (17, 1.2%) (Figure 3). Although the highest number of deaths were reported in Europe, the number of infections was also so large that Europe was the region reporting the lowest CFR. The highest CFR is seen in the Eastern Mediterranean region (5.7 deaths per 100 infections), followed by South East Asia (3.1 deaths per 100 infections) (Table 6).

**Fig 3:**
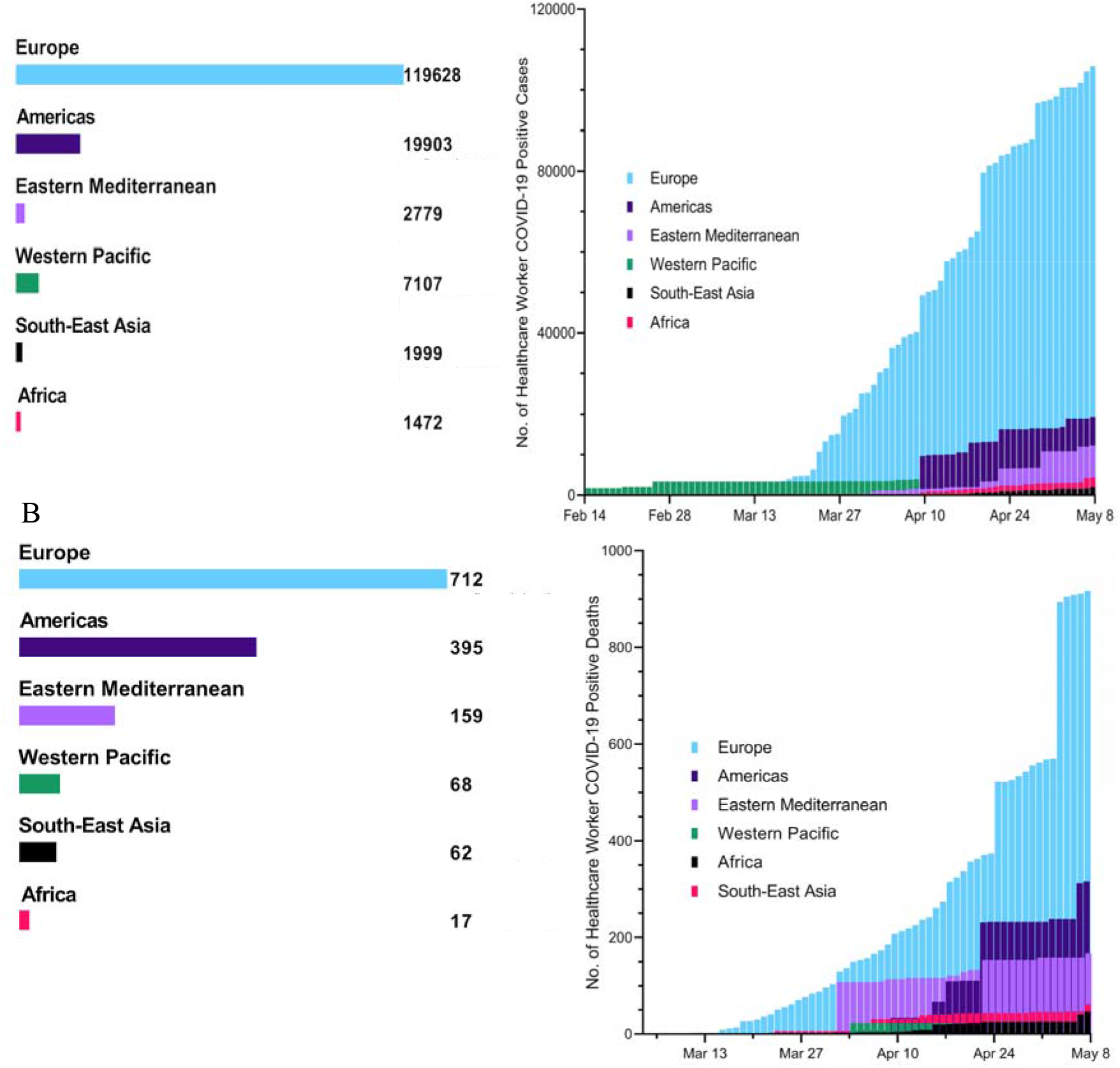
COVID-19 infections (A) & deaths (B) in healthcare workers in WHO regions.

**Table 6:** Total number of reported infections and deaths in WHO regions. *Case fatality rate is the number of reported deaths per 100 cases of reported infections.

| WHO Region | Infections | Deaths | CFR* (%) | Healthcare workers COVID-19 deaths/ Total population COVID-19 deaths (%) |
| --- | --- | --- | --- | --- |
| **Africa** | 1472 | 17 | 1.2 | 0.06 |
| **Eastern Mediterranean** | 2779 | 159 | 5.7 | 0.44 |
| **Europe** | 119628 | 712 | 0.6 | 1.40 |
| **Americas** | 19903 | 395 | 2.0 | 4.58 |
| **South-East Asia** | 1999 | 62 | 3.1 | 0.20 |
| **Western Pacific** | 7107 | 68 | 1.0 | 0.06 |
| **Total** | 152888 | 1413 | 0.92 | 0.52 |

##### 3.3.2. e Countries

On 08/05/2020, Spain reported the highest cumulative number of COVID-19 infections in HCWs in the world at 30663 (20% of all HCW infections). This is followed closely by Italy (23718) and the Netherlands (13884). Italy reported the highest cumulative number of deaths in HCWs due to COVID-19 at 220 (Figure 4). At least 10% of all COVID-19 deaths were healthcare workers deaths in 5 countries: Guyana, Venezuela, Afghanistan, Costa Rica, and Kazakhstan. Full numbers and CFRs for all countries can be found in Appendix S7 & S8. The COVID-19 infection peak varies from country to country with China and Italy demonstrating some of the earliest peaks. HCW infections and deaths reflect this as can be seen in Appendix S8. Some countries are only at the early stages of COVID-19 infection.

**Fig 4:**
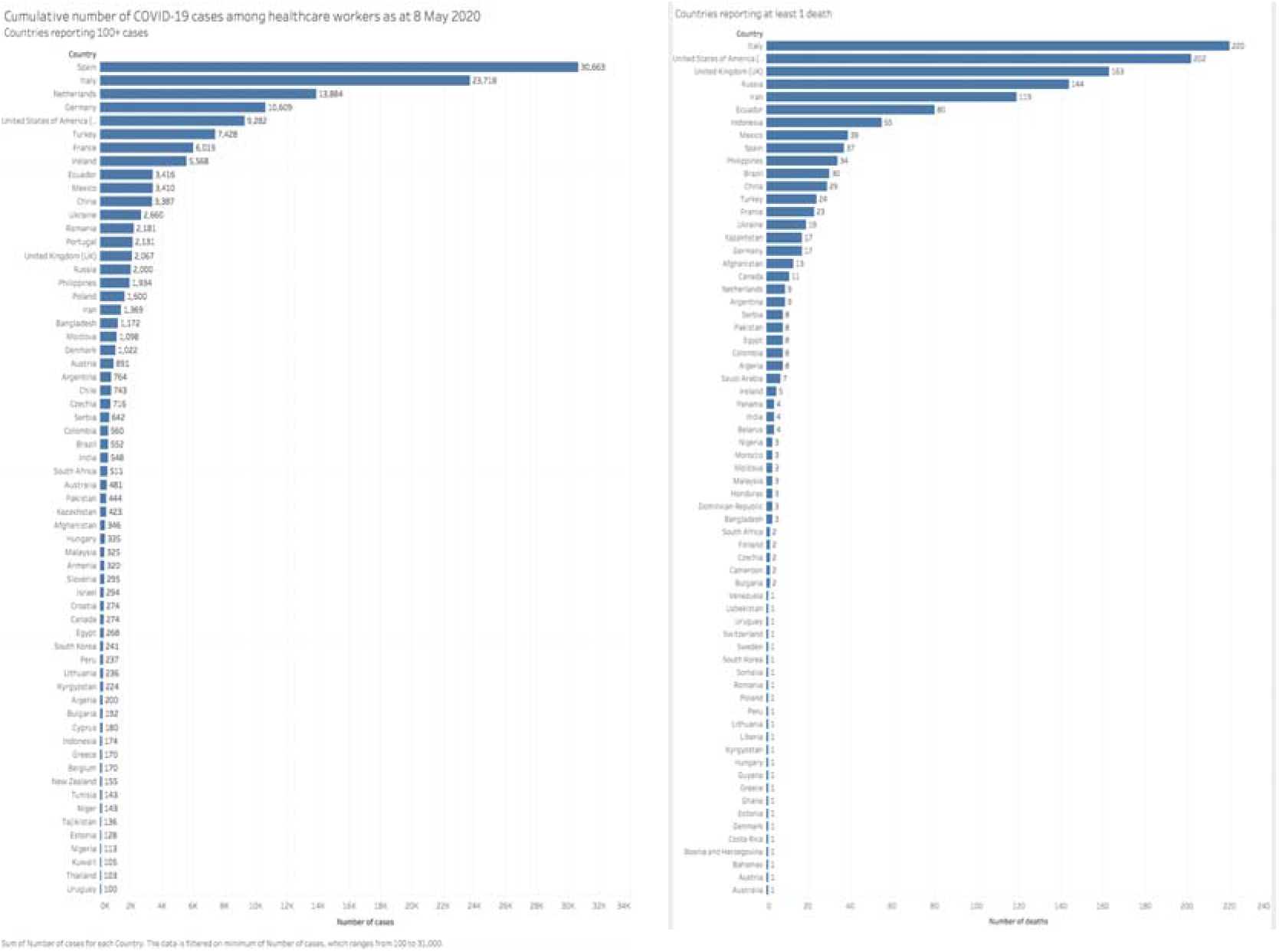
Number of reported healthcare worker infections and deaths due to COVID-19 per country up to 08/05/2020.

## 4. Discussion

Here we present the first scoping review to date examining the number of COVID-19 infections and deaths among HCWs across 195 countries. We conducted this search to estimate the infection and mortality burden among all individuals involved in the care of COVID-19 patients – from diagnosis to treatment and rehabilitation. In addition, we hoped to identify any factors associated with the risk of infection and death in HCWs.

There is an important need to address the incidence of COVID-19 related illness in HCWs globally. Failure to address infection and mortality amongst HCWs has the potential to further increase transmission of COVID-19 within healthcare facilities and their wider communities (33,34). The resulting shortage of HCWs may impair the quality of the provision of health services nationally both during the acute phase of the pandemic, and in the long term. Occupational risks in the workplace must be minimised if not altogether eliminated. Moreover, a clear pathway must be present for the early diagnosis and treatment of HCWs suspected to have contracted COVID-19. It is essential that measures are put in place to ensure that HCWs are continually protected.

### Key findings

A total of 152,888 infections and 1413 deaths were reported. Our results revealed a number of trends. The overall infection and death trends followed that of the general world population COVID-19 infection trends. Infections were mainly in women, but deaths were mainly in men. Infections were seen more in nurses, deaths more in doctors. Within the data available, GPs were the highest risk speciality for deaths amongst doctors, while the highest risk nursing speciality was mental health. It is possible that there was less PPE available in the community, with confirmed cases in hospital wards being prioritised, or that the high flow of patients through GP services has led to an increased risk of viral transmission. It may just reflect the higher number of GPs compared to hospital doctors. Mental health specialists may also be lacking PPE. Many mental health nurses also work in the community and often work in close proximity to patients, which may increase their risk of exposure. The majority of infections and deaths were reported in the 50-59 year age range, while the group aged over 70 years had the highest CFR. Europe had the highest number of infections and deaths, but the lowest CFR, while the Eastern Mediterranean had the highest CFR. By population, regions such as the Indian sub-continent and Africa reported relatively low numbers of HCW infection and death.

These trends must be considered in the context of the paucity of data, and the natural history of the disease. The differing COVID-19 infection curves in different countries is evident in the reporting trends; some countries are at the beginning of the HCW infection peak, while some are beyond. The first reported case of COVID-19 in Africa was nearly a month after the first case in Europe and Africa is slightly behind Europe in its disease course; their lower reported numbers of HCW infection is therefore unsurprising, as they are also reporting lower rates of infection overall. If this continues as the epidemic in Africa progresses, it will be necessary to consider what lessons can be learnt. Furthermore, reporting varies significantly between countries. The USA, one of the countries most severely affected by the pandemic, has not reported data about this topic for over one month. While other countries, such as the Philippines provide daily updates. The availability and quality of testing and guidelines for COVID-19 testing varies greatly across countries, which further limits the reliability of the observed trends. Estimating the percentages of HCWs infected by COVID-19 is crucial for adjusting infection prevention policies applied in the healthcare system to reduce viral transmission.

### Limitations

The primary limitation of this scoping review was the quality of the data available to us. A wide range of data was used, including grey literature, which made it difficult to normalise datasets. Furthermore, different countries were at different stages of their epidemics when we collected data. Given the incubation period of the virus before symptoms are seen and the lag in initial infection and death for those who succumb to COVID-19, data between countries at different stages of their epidemics will not be comparable. To make data comparable between different countries we would have needed to batch them according to when their epidemics started, but clear information about this was unavailable. As countries move past the peak of the virus and life begins to move back to normality, increased availability of high-quality data should allow us to conduct more extensive quantitative analysis of HCW infections. A retrospective analysis would allow countries to be matched at the same stage of the pandemic – thus allowing a like-for-like comparison.

For our primary analyses, a key limitation was the heterogeneity in HCW classification. Differences here made it difficult to accurately compare data between the countries because, for example, some countries may include all allied healthcare professionals in their numbers, others may not, which could result in reporting inaccurate proportions of HCWs infected by COVID-19. Additionally, there was limited access to accurate data about confounding variables, such as availability of testing for COVID-19 in different countries, which could have influenced infection and mortality rates among healthcare workers. Due to lack of testing, many cases of COVID-19 are diagnosed as ‘atypical pneumonia’ in some countries and thus do not feature in published figures for COVID-19 cases or deaths. Given the lack of robust data across different variables, including confounding factors, it was not possible to establish causative or even correlative links between the different variables collected and we were, therefore, limited to descriptive analyses.

### Recommendations

COVID-19 infections and deaths among HCWs necessitates provision of adequate and appropriate PPE. Infection control training must be provided for those working on the frontlines of the COVID-19 outbreak response, especially among redeployed HCWs with little experience in the clinical management of infectious diseases (35). Regulative and supportive measures must be put into place to ensure compliance with infection control policies in the workplace at all times.

The first step to achieve this would involve appropriate measures for identifying and registering those who have been infected as early as possible. Our study clearly highlights the lack of universal access to early identification measures and infection registration processes across healthcare systems in the world. The testing guidelines, access and reporting systems vary hugely across countries and are not merely a reflection of country level healthcare expenditure, although this is an important factor and further highlights inequalities between HICs and LMICs. The unavailability of relevant data in a timely manner, (which was seen in both HICs and LMICs) makes it difficult to estimate the true burden of infection and effectively plan management strategies. It also inhibits any attempt to learn from those countries beyond their peak and plan timely preventative measures in those who are yet to experience the peak. We highly recommend universal guidelines to be in place for testing and reporting of infections in HCWs in a timely manner, with consideration towards a healthcare worker international infection registry.

The gender related difference in infection and death rates in HCWs is one that has not been reported previously. Many factors may contribute towards this including more nursing staff being female and more doctors being male which may reflect differences in exposure levels, training and equipment provided, age at qualification. Further investigation of the identified trends would be recommended.

Although physicians working in certain specialities may be considered high-risk due to frequent exposure to oronasal secretions (e.g. otolaryngology, anaesthesiology, dentistry (36)), the risk to other specialities who work in other health care settings, including clinics and mental health facilities, must not be underestimated. The high rate of morbidity and mortality in elderly HCWs may require assigning them to less risky settings such as telemedicine, non-COVID-19 outpatient clinics, or administrative positions (37). HCWs who report possible symptoms and those who have had unprotected exposure to COVID-19 patients must be prioritised for testing. HCWs must be offered flexible working hours to avoid overwork, and psychological intervention plans must be implemented to help HCWs in coping with physical and psychological stress (38).

Despite its limitations, our analyses do provide a broad coverage of the data available across the world and the data we used for our analyses were run through the AACODS checklist to ensure an acceptable standard across all datasets was maintained so that we could compare them. The descriptive analyses also importantly point to the lack of reliable data in so many countries due to lack of infrastructure to quickly and robustly capture data on healthcare workers and other aspects of healthcare systems that could affect COVID-19 related morbidity and mortality among them. The countries whose datasets met the AACODS checklist criteria could serve as examples and provide best practice for countries lacking robust data collection policies and data collection systems. Our pragmatic approach in this study provides general trends to provide rapid information in response to widespread urgent calls from healthcare workers worldwide.

## Data Availability

All data is available in the manuscript and appendices

## Declarations

### Author contributions

RK is the guarantor. RK and SB conceived the project. SB, RK, and AJ contributed equally to the design of the project. All authors contributed to the collection of the data. SB, REB, MA, DO, YB, AK, SAP, GB, DK, SC, MK, AJ, AM and RK drafted the manuscript. All remaining authors extensively reviewed the manuscript. All authors have approved the final manuscript and are willing to take responsibility for appropriate portions of the content. The corresponding author attests that all listed authors meet authorship criteria and that no others meeting the criteria have been omitted.

## Acknowledgements

Kokila Lakhoo and Eve Thangaraj for their advice.

## Appendix

### Appendix S1: The Protocol

#### Review question(s)

Our primary aim is to perform a scoping review to estimate the number and proportion of health care workers who have become infected with COVID-19 in every country in the world.

Our secondary aims are:

1. to establish health care worker mortality rate linked to COVID-19 in every country in the world, and
2. to identify factors that could be linked to levels of infection and mortality of health care workers between countries.

#### Searches

A full systematic search of bibliographic databases will be performed - Embase and MEDLINE. All databases will be searched from 17^th^ November 2019 to 22^nd^ April 2020 without language restriction for all terms related to health care workers and COVID-19 (Appendix S1). The search results will be merged, and duplicate citations will be discarded. Titles and abstracts will be screened by two reviewers independently based on the pre-defined inclusion and exclusion criteria. The full text of the remaining articles will be retrieved and screened by two reviewers independently. Conflicts are to be resolved by mutual agreement or by a third reviewer. The reference lists of included documents will be examined to identify any further relevant documents missed through the above search strategy.

A grey literature search of WHO documents, government documents, and non-governmental organisation documents will also be conducted. Social media sites, media websites, and google will be utilised to find these documents and cross-reference sources. All documents will be collected by two reviewers independently. The reference lists of included documents will be examined to identify any further relevant documents missed through the above search strategy. The inclusion of the documents and data extracted from them will be compared between the two reviewers and validated by a third reviewer.

The search strategy outlined above will be performed by individuals who have experience in research methodologies. All reviewers will attend an online training and support session delivered by SB and RK before performing any searches. The Preferred Reporting Items for Systematic Reviews and Meta-Analysis extension for Scoping Reviews (PRISMA-ScR) guidelines will be used to write and report the findings

#### Inclusion Criteria

##### Types of study to be included

All studies, synopses of studies, synthesis, synopses of synthesis, and summaries are eligible to be included. Primary data – where available – will be eligible for inclusion

Studies will be excluded if they do not use real human data or do not state their methodology.

##### Condition(s) or domain(s) to be included

The infection and mortality of health care workers associated with COVID-19 in all country settings. For the purposes of this review, a country is that which is recognised by the United Nations (UN) to be a sovereign country.

##### Participants/population to be included

Inclusion:

- Health care workers. For the purposes of this review, a health care worker is one who delivers care and services to the sick and ailing either directly as paramedics, nurses and doctors, or indirectly as aides, helpers, laboratory technicians, and medical waste handlers.

Exclusion:

- Animal studies
- Statistical modelling

##### Intervention(s), exposure(s)

COVID-19 in a health care worker

##### Comparator(s)/control

Not applicable

##### Context

This review includes settings at all levels of development. It considers low-, middle-, and high-socio-demographic index (SDI) countries

##### Main outcome(s)

1. COVID 19 infections in healthcare workers (a) worldwide and (b) by country
2. Healthcare workers deaths related to COVID 19 (could be with or from) (a) worldwide and (b) by country

##### *Measures of effect

There is no restriction on time to mortality outcome

##### Additional outcome(s)

Demographics of health care workers who have been (a) infected with and (b) died from COVID-19 Factors that could be linked to infection and mortality of health care workers with COVID-19

##### *Measures of effect

Not applicable

##### Data extraction (selection and coding)

Using a pre-designed and pre-piloted data extraction form, data from each included document will be collected by two independent reviewers. Conflicts in data collection will be resolved by a third reviewer. From all included documents information will be extracted on study design, study setting, study population, participant demographics, timeframe of the study, date of publication, public health measures implemented, health care worker infected with COVID-19, health care worker mortality related to COVID-19, and information for assessment of the risk of bias.

##### Risk of bias (quality) assessment

Two reviewers will independently classify the risk of bias in each included document using the risk of bias in randomised trials 2 (ROB 2) tool, risk of bias in non-randomised studies (ROBINS I) tool, assessing the methodological quality of systematic reviews (AMSTAR) tool, and AACODS checklist as appropriate. Documents will be graded as high (bias is very likely due to essential errors), moderate (no essential deficiencies, but not all criteria are met), low (bias is unlikely), or unclear.

The reviewers will discuss and resolve any disagreements with the level of bias in a study.

##### Strategy for data synthesis

A random effects model will be used to pool mortality and infection rates. Where possible, associations will be analysed by computing and/or pooling this estimation using a random effects meta-analysis.

Health care workforce deaths due to COVID-19 as a proportion of total population deaths due to COVID-19 will be calculated. Health care workforce deaths due to COVID-19 as a proportion of all health care work force COVID-19 infections and health care workforce deaths due to COVID-19 as a proportion of the total healthcare workforce will be calculated and compared to publicly available total population data. Prevalence and risk ratios will be given.

##### Analysis of subgroups or subsets

Where possible, subgroup analysis will be by region (by world bank), SDI status (low SDI country, middle SDI country, and high SDI country), age range (18-29; 30-39; 40-49; 50-59; 60-70), gender (male/female), ethnicity, type of healthcare worker, sub-specialities, and level of training.

### Appendix S2: PRISMA – ScR Checklist

| SECTION | ITEM | PRISMA-ScR CHECKLIST ITEM | REPORTED ON PAGE # |
| --- | --- | --- | --- |
| **TITLE** |  |  |  |
| Title | 1 | Identify the report as a scoping review. | 1 |
| **ABSTRACT** |  |  |  |
| Structured summary | 2 | Provide a structured summary that includes (as applicable): background, objectives, eligibility criteria, sources of evidence, charting methods, results, and conclusions that relate to the review questions and objectives. | 2 |
| **INTRODUCTION** |  |  |  |
| Rationale | 3 | Describe the rationale for the review in the context of what is already known. Explain why the review questions/objectives lend themselves to a scoping review approach. | 3 |
| Objectives | 4 | Provide an explicit statement of the questions and objectives being addressed with reference to their key elements (e.g., population or participants, concepts, and context) or other relevant key elements used to conceptualize the review questions and/or objectives. | 4 |
| **METHODS** |  |  |  |
| Protocol and registration | 5 | Indicate whether a review protocol exists; state if and where it can be accessed (e.g., a Web address); and if available, provide registration information, including the registration number. | 4 |
| Eligibility criteria | 6 | Specify characteristics of the sources of evidence used as eligibility criteria (e.g., years considered, language, and publication status), and provide a rationale. | 4 |
| Information sources* | 7 | Describe all information sources in the search (e.g., databases with dates of coverage and contact with authors to identify additional sources), as well as the date the most recent search was executed. | 4-5 |
| Search | 8 | Present the full electronic search strategy for at least 1 database, including any limits used, such that it could be repeated. | 4 |
| Selection of sources of evidence† | 9 | State the process for selecting sources of evidence (i.e., screening and eligibility) included in the scoping review. | 5 |
| Data charting process‡ | 10 | Describe the methods of charting data from the included sources of evidence (e.g., calibrated forms or forms that have been tested by the team before their use, and whether data charting was done independently or in duplicate) and any processes for obtaining and confirming data from investigators. | 5 |
| Data items | 11 | List and define all variables for which data were sought and any assumptions and simplifications made. | 5 |
| Critical appraisal of individual sources of evidence§ | 12 | If done, provide a rationale for conducting a critical appraisal of included sources of evidence; describe the methods used and how this information was used in any data synthesis (if appropriate). | 6 |
| Synthesis of results | 13 | Describe the methods of handling and summarizing the data that were charted. | 6 |
| **RESULTS** |  |  |  |
| Selection of sources of evidence | 14 | Give numbers of sources of evidence screened, assessed for eligibility, and included in the review, with reasons for exclusions at each stage, ideally using a flow diagram. | 6-7 |
| Characteristics of sources of evidence | 15 | For each source of evidence, present characteristics for which data were charted and provide the citations. | 7-8 |
| Critical appraisal within sources of evidence | 16 | If done, present data on critical appraisal of included sources of evidence (see item 12). | 25 |
| Results of individual sources of evidence | 17 | For each included source of evidence, present the relevant data that were charted that relate to the review questions and objectives. | 8-15 |
| Synthesis of results | 18 | Summarize and/or present the charting results as they relate to the review questions and objectives. | 15-16 |
| **DISCUSSION** |  |  |  |
| Summary of evidence | 19 | Summarize the main results (including an overview of concepts, themes, and types of evidence available), link to the review questions and objectives, and consider the relevance to key groups. | 15-16 |
| Limitations | 20 | Discuss the limitations of the scoping review process. | 16 |
| Conclusions | 21 | Provide a general interpretation of the results with respect to the review questions and objectives, as well as potential implications and/or next steps. | 17 |
| **FUNDING** |  |  |  |
| Funding | 22 | Describe sources of funding for the included sources of evidence, as well as sources of funding for the scoping review. Describe the role of the funders of the scoping review. | 17 |

### Appendix S3: The Search Terms Used

- health care practitioner
- health care professional
- health care provider
- health care worker
- health personnel
- health profession personnel
- health worker
- healthcare personnel
- healthcare practitioner
- healthcare professional
- healthcare provider
- healthcare worker
- health care manpower
- health labour force
- health manpower
- health work force
- healthcare labor force
- healthcare labour force
- healthcare manpower
- healthcare work force
- healthcare workforce
- medical manpower
- doctor
- medical practitioner
- physician associate
- physicians
- practitioner
- private physician
- nurse
- nurses’ aides
- nursing aid
- nursing aide
- nursing assistants
- orderlies
- porters
- healthcare assistants
- physician assistant
- advanced clinical practitioner
- advanced practice clinician
- advanced practice professional
- allied health provider
- clinical associate
- limited-license practitioner
- mid-level practitioner
- mid-level provider
- non-physician practitioner
- non-physician provider
- physician extender
- care coordinator
- health care coordinator
- healthcare coordinator
- medical dispatcher
- accredited social health activist
- ASHA (accredited social health activist)
- ASHA workers
- auxiliary health worker
- barefoot doctor
- health practitioner
- health aides
- health officers
- medical chaperone
- medical expert
- medical staff
- physician assistant
- psychotherapist
- physiotherapist
- occupational therapist
- pharmacist
- allied health personnel
- paramedical personnel
- para medical personnel
- paramedical assistant
- paramedical manpower
- paramedical professional
- paramedical staff
- psychiatric aides
- medical student
- student nurse
- corona
- coronavirus
- COVID
- COVID-19
- SARS-CoV-2
- Pandemic

The search was restricted to 2019 – 2020, and humans.

### Appendix S4: Data Sources and The Quality of These SourcesPercent of Sources (%)

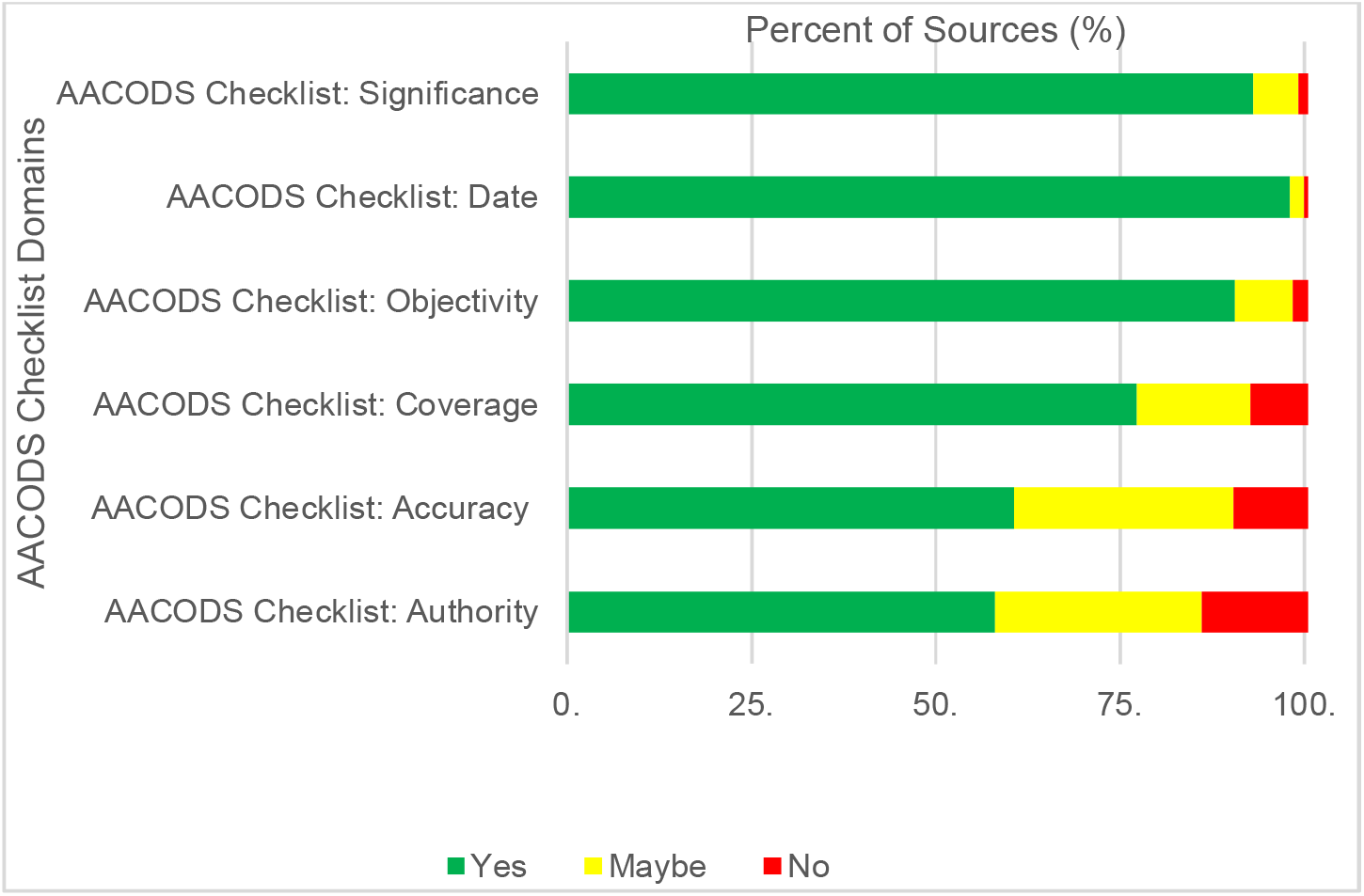
*Fig. Appraisal of sources* Please see Excel Spreadsheet entitled ‘Data Sources and The Quality of These Sources’

### Appendix S5: Data Extraction

Please see Excel Spreadsheet entitled ‘Data Extraction’

### Appendix S6: AACODS Checklist

| AACODS |  | YES | NO | ? |
| --- | --- | --- | --- | --- |
| **Authority** | Identifying who is responsible for the intellectual content.<br>**Individual author:**<br>Associated with a reputable organisation?<br>Professional qualifications or considerable experience?<br>Produced/published other work (grey/black) in the field?<br>Recognised expert, identified in other sources?<br>Cited by others? (use Google Scholar as a quick check)<br>Higher degree student under "expert" supervision?<br>**Organisation or group:**<br>Is the organisation reputable? (e.g. W.H.O)<br>Is the organisation an authority in the field?<br>**In all cases:**<br>Does the item have a detailed reference list or bibliography? |  |  |  |
| **Accuracy** | Does the item have a clearly stated aim or brief?<br>Is so, is this met?<br>Does it have a stated methodology?<br>If so, is it adhered to?<br>Has it been peer-reviewed?<br>Has it been edited by a reputable authority?<br>Supported by authoritative, documented references or credible sources?<br>Is it representative of work in the field?<br>If No, is it a valid counterbalance?<br>Is any data collection explicit and appropriate for the research?<br>If item is secondary material (e.g. a policy brief of a technical report) refer to the original. Is it an accurate, unbiased interpretation or analysis? |  |  |  |
| **Coverage** | All items have parameters which define their content coverage. These limits might mean that a work refers to a particular population group, or that it excluded certain types of publication. A report could be designed to answer a particular question, or be based on statistics from a particular survey.<br>• Are any limits clearly stated? |  |  |  |
| **Objectivity** | It is important to identify bias, particularly if it is unstated or unacknowledged.<br>• Opinion, expert or otherwise, is still opinion: is the author's standpoint clear?<br>• Does the work seem to be balanced in presentation? |  |  |  |
| **Date** | For the item to inform your research, it needs to have a date that confirms relevance<br>• Does the item have a clearly stated date related to content? No easily discernible date is a strong concern.<br>• If no date is given, but can be closely ascertained, is there a valid reason for its absence?<br>• Check the bibliography: have key contemporary material been included? |  |  |  |
| **Significance** | This is a value judgment of the item, in the context of the relevant research area<br>• Is the item meaningful? (this incorporates feasibility, utility and relevance)<br>• Does it add context?<br>• Does it enrich or add something unique to the research?<br>• Does it strengthen or refute a current position?<br>• Would the research area be lesser without it?<br>• Is it integral, representative, typical?<br>• Does it have impact? (in the sense of influencing the work or behaviour of others) |  |  |  |

### Appendix S7: Supplementary figures for the number of COVID-19 infections and deaths reported in healthcare worker per country as of 08/05/2020

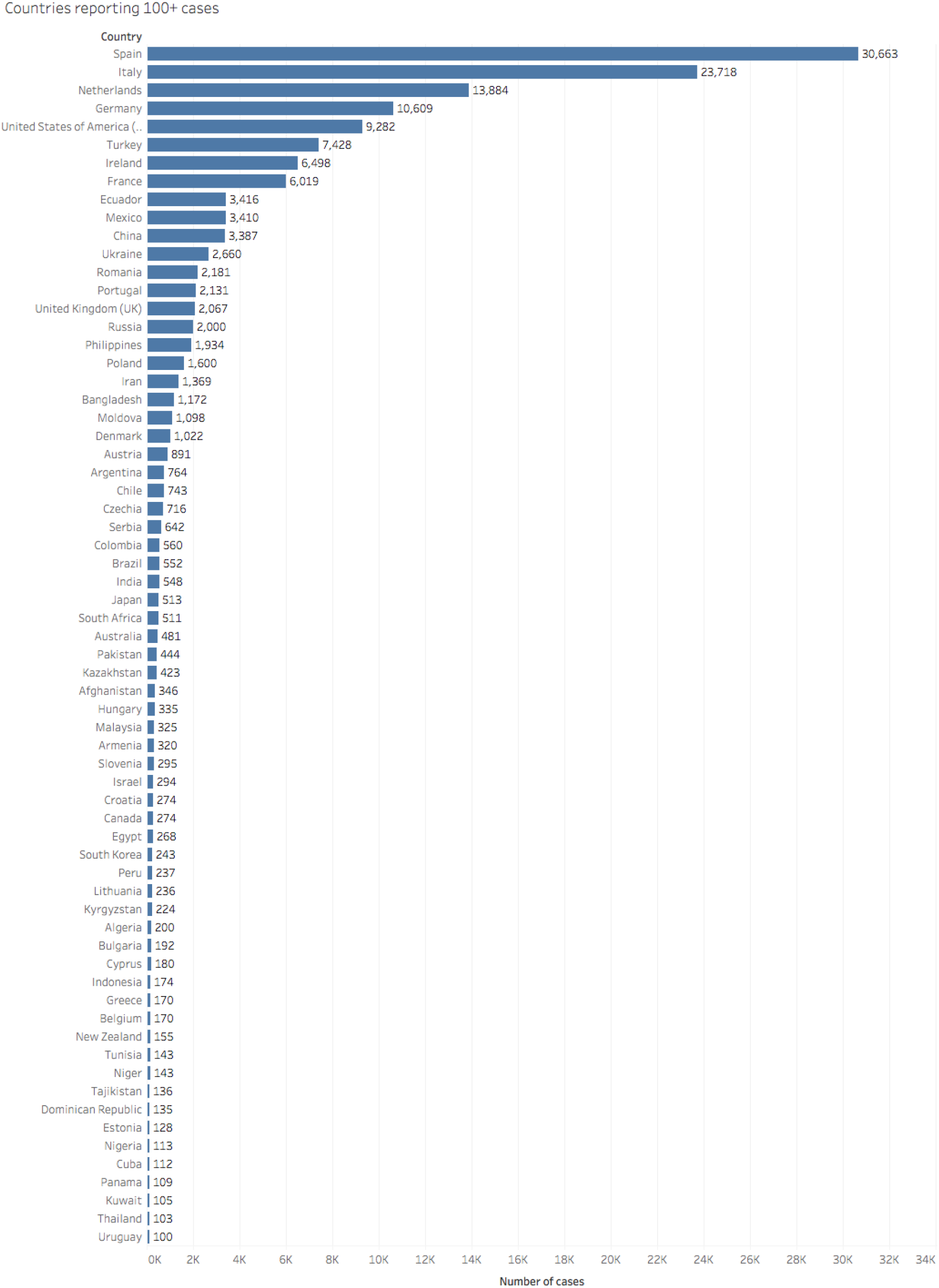
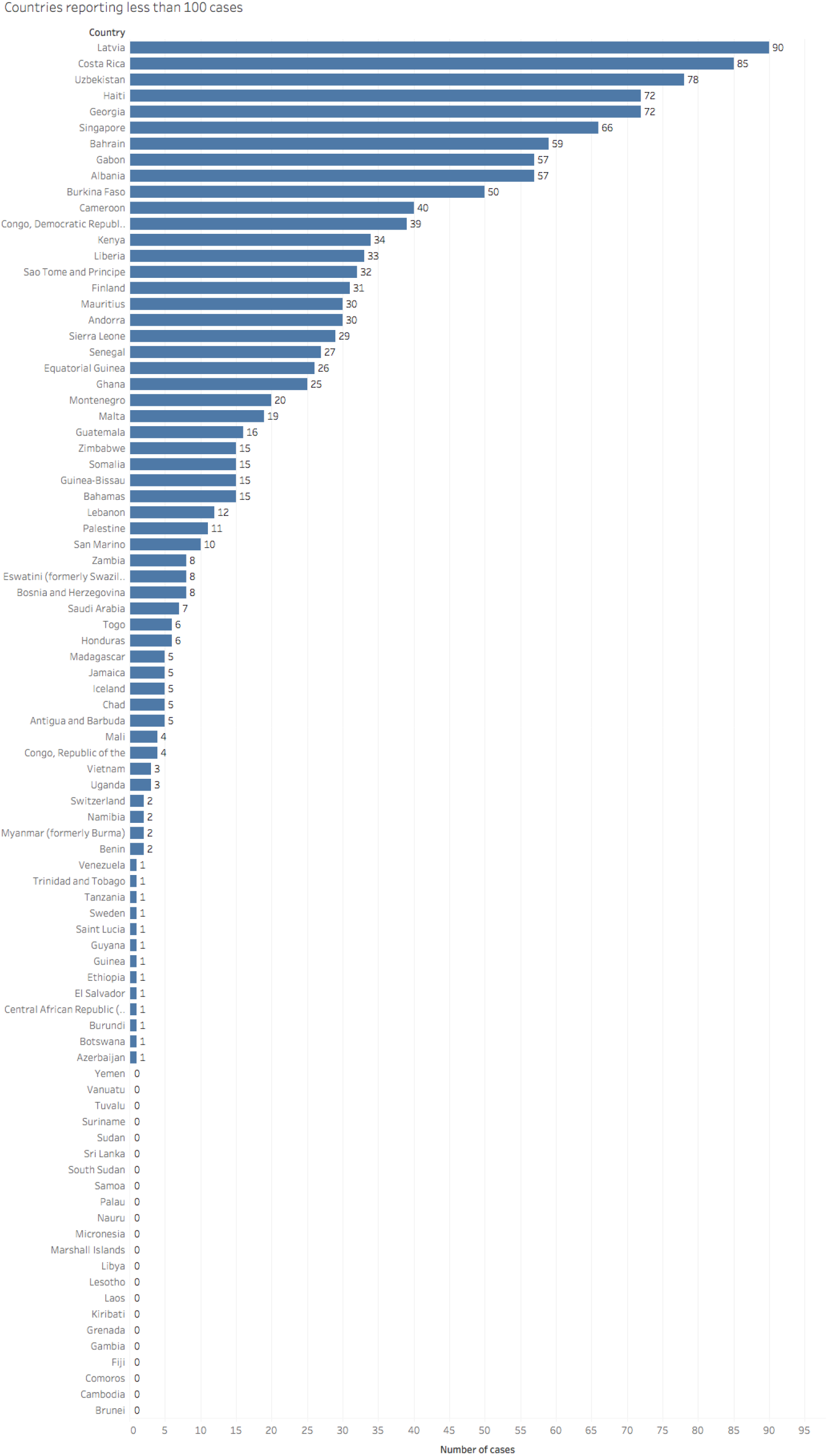
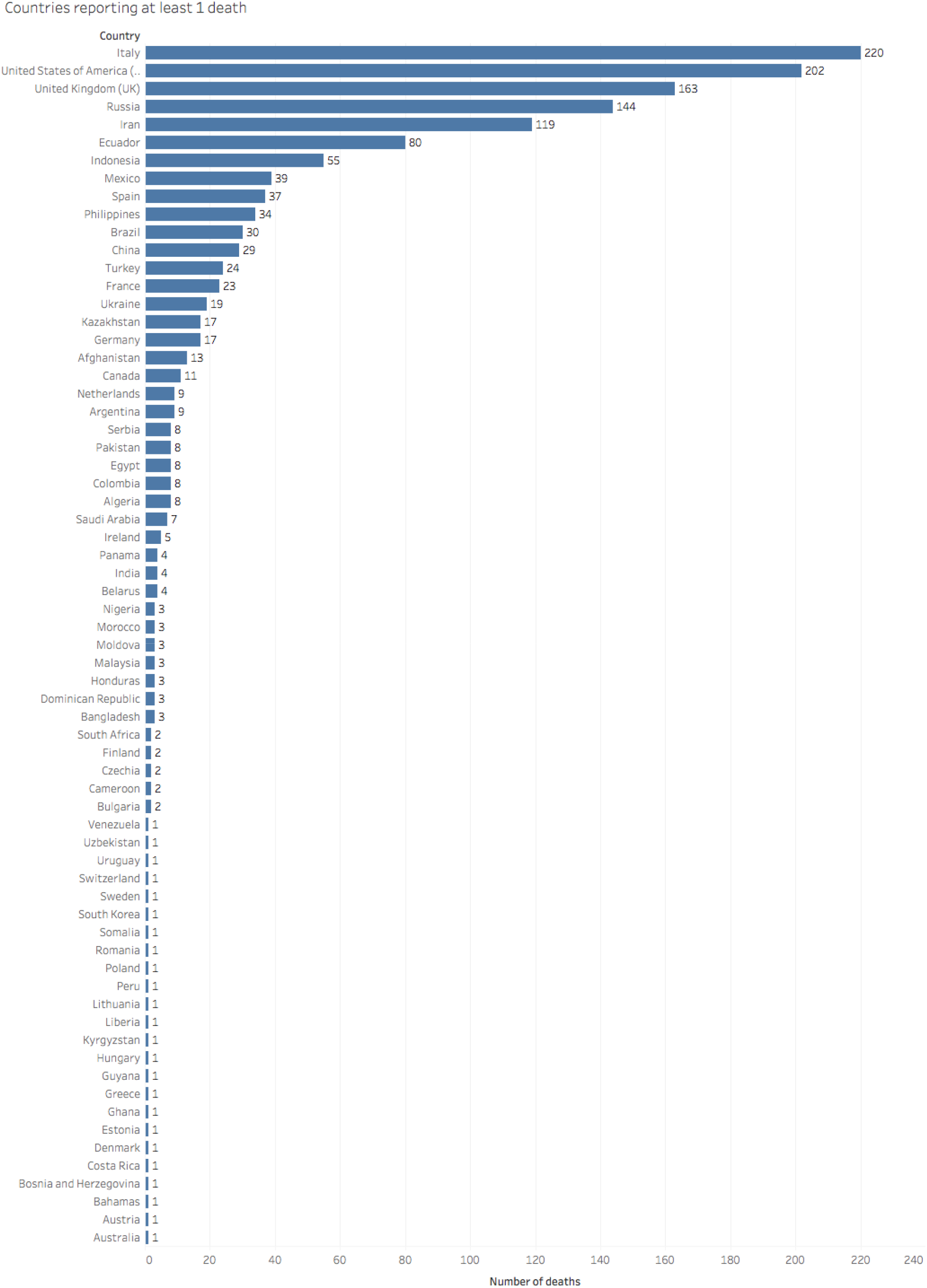

| Name of country | COVID-19 deaths among healthcare workers | COVID-19 cases among healthcare workers | CFR (Deaths as Percentage of total HCW infections) | Death as percentage of total population COVID deaths | Infections as percentage of total tests (%) |
| --- | --- | --- | --- | --- | --- |
| **Afghanistan** | 13 | 346 | 3.8 | 12.3 | 5.4 |
| **Albania** | 0 | 57 | 0.0 | 0.0 | 0.8 |
| **Algeria** | 8 | 200 | 4.0 | 1.7 | 6.7 |
| **Andorra** | 0 | 30 | 0.0 | 0.0 | 1.8 |
| **Angola** | Unreported |  |  |  |  |
| **Antigua and Barbuda** | 0 | 5 | 0.0 | 0.0 | 4.6 |
| **Argentina** | 9 | 764 | 1.2 | 3.3 | 1.3 |
| **Armenia** | 0 | 320 | 0.0 | 0.0 | 20.2 |
| **Australia** | 1 | 481 | 0.2 | 1.0 | 0.1 |
| **Austria** | 1 | 891 | 0.1 | 0.2 | 0.4 |
| **Azerbaijan** | 0 | 1 | 0.0 | 0.0 | <0.1 |
| **Bahamas** | 1 | 15 | 6.7 | 9.1 |  |
| **Bahrain** | 0 | 59 | 0.0 | 0.0 | 0.0 |
| **Bangladesh** | 3 | 1172 | 0.3 | 1.5 | 1.1 |
| **Barbados** | Unreported |  |  |  |  |
| **Belarus** | 4 | 0 | N/A | 3.4 | 0.0 |
| **Belgium** | 0 | 170 | 0.0 | 0.0 | <0.1 |
| **Belize** | Unreported |  |  |  |  |
| **Benin** | 0 | 2 | 0.0 | 0.0 | <0.1 |
| **Bhutan** | Unreported |  |  |  |  |
| **Bolivia** | Unreported |  |  |  |  |
| **Bosnia and Herzegovina** | 1 | 8 | 12.5 | 1.1 |  |
| **Botswana** | 0 | 1 | 0.0 | 0.0 | <0.1 |
| **Brazil** | 30 | 552 | 5.4 | 0.3 | 0.2 |
| **Brunei** | Unreported |  |  |  |  |
| **Bulgaria** | 2 | 192 | 1.0 | 2.4 | 0.4 |
| **Burkina Faso** | 0 | 50 | 0.0 | 0.0 |  |
| **Burundi** | 0 | 1 | 0.0 | 0.0 | <0.1 |
| **Cabo Verde** | Unreported |  |  |  |  |
| **Cambodia** | Unreported |  |  |  |  |
| **Cameroon** | 2 | 40 | 5.0 | 1.9 |  |
| **Canada** | 11 | 274 | 4.0 | 0.2 | <0.1 |
| **Central African Republic (CAR)** | 0 | 1 | 0.0 | N/A | <0.1 |
| **Chad** | 0 | 5 | 0.0 | 0.0 |  |
| **Chile** | 0 | 743 | 0.0 | 0.0 | 0.3 |
| **China** | 29 | 3,387 | 0.9 | 0.6 |  |
| **Colombia** | 8 | 560 | 1.4 | 2.0 | 0.4 |
| **Comoros** | Unreported |  |  |  |  |
| **Congo, Democratic Republic of the** | 0 | 39 | 0.0 | 0.0 |  |
| **Congo, Republic of the** | 0 | 4 | 0.0 | 0.0 |  |
| **Costa Rica** | 1 | 85 | 1.2 | 16.7 | 0.7 |
| **Cote d'Ivoire** | Unreported |  |  |  |  |
| **Croatia** | 0 | 274 | 0.0 | 0.0 | 0.7 |
| **Cuba** | 0 | 92 | 0.0 | 0.0 | 0.2 |
| **Cyprus** | 0 | 180 | 0.0 | 0.0 | 0.5 |
| **Czechia** | 2 | 716 | 0.3 | 0.7 | 0.2 |
| **Denmark** | 1 | 1022 | 0.1 | 0.2 | 0.4 |
| **Djibouti** | Unreported |  |  |  |  |
| **Dominica** | Unreported |  |  |  |  |
| **Dominican Republic** | 3 | 135 | 2.2 |  | 0.7 |
| **Ecuador** | 80 | 3416 | 2.3 | 4.8 | 5.4 |
| **Egypt** | 8 | 268 | 3.0 | 1.7 | 0.5 |
| **El Salvador** | 0 | 1 | 0.0 | 0.0 | <0.1 |
| **Equatorial Guinea** | 0 | 26 | 0.0 | 0.0 | 3.0 |
| **Eritrea** | Unreported |  |  |  |  |
| **Estonia** | 1 | 128 | 0.8 | 1.8 | 0.3 |
| **Eswatini (formerly Swaziland)** | 0 | 8 | 0.0 | 0.0 | 0.8 |
| **Ethiopia** | 0 | 1 | 0.0 | 0.0 | <0.1 |
| **Fiji** | Unreported |  |  |  |  |
| **Finland** | 2 | 31 | 6.5 | 0.8 | <0.1 |
| **France** | 23 | 6019 | 0.4 | 0.1 | 0.7 |
| **Gabon** | 0 | 57 | 0.0 | 0.0 | 10.0 |
| **Gambia** | Unreported |  |  |  |  |
| **Georgia** | 0 | 72 | 0.0 | 0.0 |  |
| **Germany** | 17 | 10609 | 0.2 | 0.2 | 0.4 |
| **Ghana** | 1 | 25 | 4.0 | 5.6 | <0.1 |
| **Greece** | 0 | 170 | 0.0 | 0.0 | 0.2 |
| **Grenada** | Unreported |  |  |  |  |
| **Guatemala** | 0 | 16 | 0.0 | 0.0 | 0.2 |
| **Guinea** | 0 | 1 | 0.0 | 0.0 | <0.1 |
| **Guinea-Bissau** | 0 | 15 | 0.0 | 0.0 | 1.0 |
| **Guyana** | 1 | Unreported | N/A | 10.0 | 0.3 |
| **Haiti** | 0 | 72 | 0.0 | 0.0 |  |
| **Honduras** | 3 | 6 | 50.0 | 3.0 |  |
| **Hungary** | 1 | 335 | 0.3 | 0.3 | 0.3 |
| **Iceland** | 0 | 5 | 0.0 | 0.0 | <0.1 |
| **India** | 4 | 548 | 0.5 | 0.2 | <0.1 |
| **Indonesia** | 55 | 174 | 31.6 | 5.9 | 0.2 |
| **Iran** | 119 | 1369 | 8.7 | 1.8 | 0.3 |
| **Iraq** | Unreported |  |  |  |  |
| **Ireland** | 5 | 5568 | 0.1 | 0.4 | 3.0 |
| **Israel** | 0 | 294 | 0.0 | 0.0 | 0.1 |
| **Italy** | 220 | 23718 | 0.9 | 0.7 | 1.0 |
| **Jamaica** | 0 | 5 | 0.0 | 0.0 | 0.2 |
| **Japan** | 0 | 44 | 0.0 | 0.0 | 0.3 |
| **Jordan** | Unreported |  |  |  |  |
| **Kazakhstan** | 17 | 423 | 4.0 | 56.7 | 0.1 |
| **Kenya** | 0 | 10 | 0.0 | 0.0 | 0.1 |
| **Kiribati** | Unreported |  |  |  |  |
| **Kuwait** | 0 | 105 | 0.0 | 0.0 |  |
| **Kyrgyzstan** | 1 | 224 | 0.4 | 8.3 | 0.7 |
| **Laos** | Unreported |  |  |  |  |
| **Latvia** | 0 | 90 | 0.0 | 0.0 | 0.1 |
| **Lebanon** | 0 | 12 | 0.0 | 0.0 |  |
| **Lesotho** | Unreported |  |  |  |  |
| **Liberia** | 1 | 33 | 3.0 | 5.0 |  |
| **Libya** | Unreported |  |  |  |  |
| **Liechtenstein** | Unreported |  |  |  |  |
| **Lithuania** | 1 | 236 | 0.4 | 2.0 | 0.1 |
| **Luxembourg** | Unreported |  |  |  |  |
| **Madagascar** | 0 | 5 | 0.0 | N/A | 0.2 |
| **Malawi** | Unreported |  |  |  |  |
| **Malaysia** | 3 | 325 | 0.9 | 2.8 | 0.1 |
| **Maldives** | Unreported |  |  |  |  |
| **Mali** | 0 | 4 | 0.0 | 0.0 | 0.2 |
| **Malta** | 0 | 19 | 0.0 | 0.0 | <0.1 |
| **Marshall Islands** | Unreported |  |  |  |  |
| **Mauritania** | Unreported |  |  |  |  |
| **Mauritius** | 0 | 30 | 0.0 | 0.0 | 0.3 |
| **Mexico** | 39 | 3410 | 1.1 | 1.4 | 3.5 |
| **Micronesia** | Unreported |  |  |  |  |
| **Moldova** | 3 | 1098 | 0.3 | 2.1 |  |
| **Monaco** | Unreported |  |  |  |  |
| **Mongolia** | Unreported |  |  |  |  |
| **Montenegro** | 0 | 20 | 0.0 | 0.0 | 0.4 |
| **Morocco** | 3 | 0 | N/A | 1.6 | 0 |
| **Mozambique** | Unreported |  |  |  |  |
| **Myanmar (formerly Burma)** | 0 | 1 | 0.0 | 0.0 | <0.1 |
| **Namibia** | 0 | 2 | 0.0 | N/A | 0.4 |
| **Nauru** | Unreported |  |  |  |  |
| **Nepal** | Unreported |  |  |  |  |
| **Netherlands** | 9 | 13884 | 0.1 | 0.2 | 5.6 |
| **New Zealand** | 0 | 155 | 0.0 | 0.0 | <0.1 |
| **Nicaragua** | Unreported |  |  |  |  |
| **Niger** | 0 | 143 | 0.0 | 0.0 |  |
| **Nigeria** | 3 | 113 | 2.7 | 2.9 | 0.5 |
| **North Korea** | Unreported |  |  |  |  |
| **North Macedonia (formerly Macedonia)** | Unreported |  |  |  |  |
| **Norway** | Unreported |  |  |  |  |
| **Oman** | Unreported |  |  |  |  |
| **Pakistan** | 8 | 444 | 1.8 | 1.4 | 0.2 |
| **Palau** | Unreported |  |  |  |  |
| **Palestine** | 0 | 11 | 0.0 | 0.0 | <0.1 |
| **Panama** | 4 | 0 | N/A | 1.8 | 0.3 |
| **Papua New Guinea** | Unreported |  |  |  |  |
| **Paraguay** | Unreported |  |  |  |  |
| **Peru** | 1 | 237 | 0.4 | 0.1 | <0.1 |
| **Philippines** | 34 | 0 | N/A | 5.0 | 1.5 |
| **Poland** | 1 | 1600 | 0.1 | 0.1 | 0.4 |
| **Portugal** | 0 | 2131 | 0.0 | 0.0 | 0.4 |
| **Qatar** | Unreported |  |  |  | 0 |
| **Romania** | 1 | 2181 | 0.0 | 0.1 | 1.0 |
| **Russia** | 144 | 2000 | 7.2 | 8.9 | <0.1 |
| **Rwanda** | Unreported |  |  |  |  |
| **Saint Kitts and Nevis** | Unreported |  |  |  |  |
| **Saint Lucia** | 0 | 1 | 0.0 | N/A | 0.3 |
| **Saint Vincent and the Grenadines** | Unreported |  |  |  |  |
| **Samoa** | Unreported |  |  |  |  |
| **San Marino** | 0 | 10 | 0.0 | 0.0 | 0.5 |
| **Sao Tome and Principe** | 0 | 32 | 0.0 | 0.0 |  |
| **Saudi Arabia** | 7 | 5 | 140.0 | 3.2 | <0.1 |
| **Senegal** | 0 | 27 | 0.0 | 0.0 | 0.1 |
| **Serbia** | 8 | 642 | 1.2 | 3.9 | 0.5 |
| **Seychelles** | Unreported |  |  |  |  |
| **Sierra Leone** | 0 | 29 | 0.0 | 0.0 |  |
| **Singapore** | 0 | 66 | 0.0 | 0.0 | <0.1 |
| **Slovakia** | Unreported |  |  |  |  |
| **Slovenia** | 0 | 295 | 0.0 | 0.0 | 0.5 |
| **Solomon Islands** | Unreported |  |  |  |  |
| **Somalia** | 1 | 6 | 16.7 | 2.3 | 6.3 |
| **South Africa** | 2 | 511 | 0.4 | 1.2 | 0.2 |
| **South Korea** | 1 | 241 | 0.4 | 0.4 | <0.1 |
| **South Sudan** | Unreported |  |  |  |  |
| **Spain** | 37 | 30663 | 0.1 | 0.1 | 2.3 |
| **Sri Lanka** | Unreported |  |  |  |  |
| **Sudan** | Unreported |  |  |  |  |
| **Suriname** | Unreported |  |  |  |  |
| **Sweden** | 1 | 0 | N/A | 0.0 |  |
| **Switzerland** | Unreported |  |  |  | <0.1 |
| **Syria** | Unreported |  |  |  |  |
| **Tajikistan** | 0 | 136 | 0.0 | 0.0 |  |
| **Tanzania** | 0 | 1 | 0.0 | 0.0 |  |
| **Thailand** | 0 | 103 | 0.0 | 0.0 | 0.1 |
| **Timor-Leste** | Unreported |  |  |  |  |
| **Togo** | 0 | 6 | 0.0 | 0.0 | 0.1 |
| **Tonga** | Unreported |  |  |  |  |
| **Trinidad and Tobago** | 0 | 1 | 0.0 | 0.0 | 0.1 |
| **Tunisia** | 0 | 143 | 0.0 | 0.0 | 0.6 |

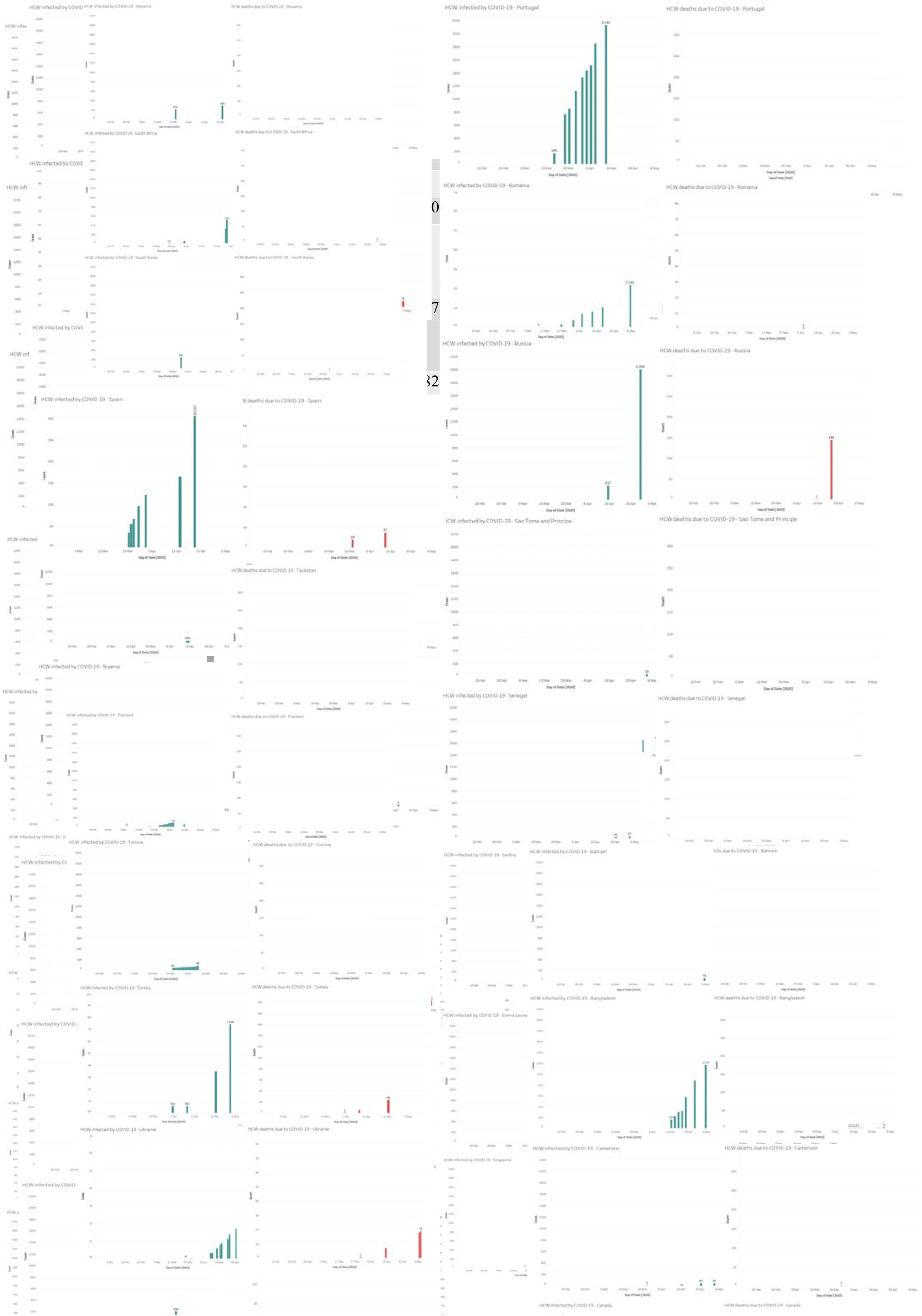

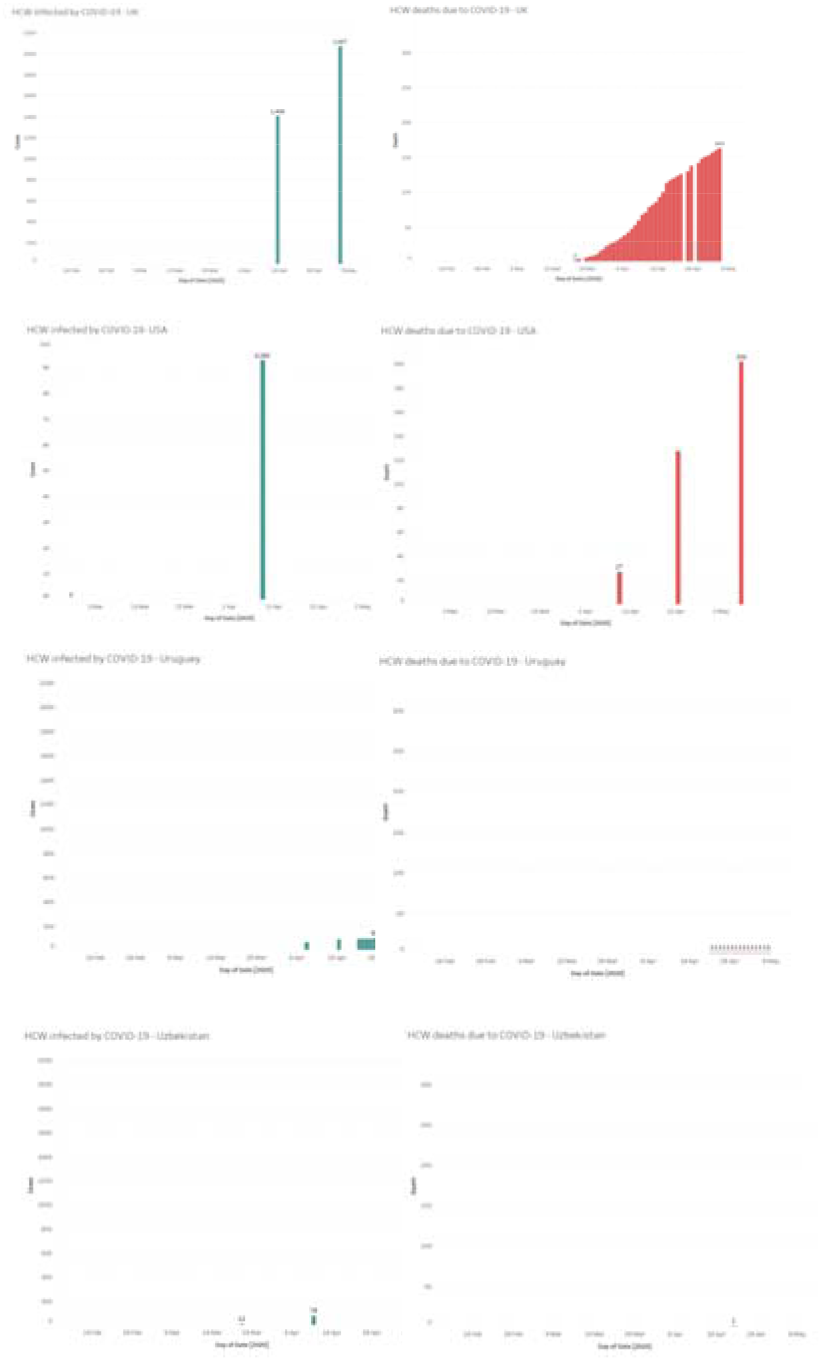

## References

1. World Health Organization. WHO Director-General’s opening remarks at the media briefing on COVID-19 - 11 March 2020 [Internet]. [cited 2020 May 3]. Available from: https://www.who.int/dg/speeches/detail/who-director-general-s-opening-remarks-at-the-media-briefing-on-covid-19---11-march-2020

2. Zhu N, Zhang D, Wang W, Li X, Yang B, Song J, et al. A novel coronavirus from patients with pneumonia in China, 2019. N Engl J Med. 2020 Feb 20;382(8):727–33.

3. World Health Organization. Coronavirus disease 2019 [Internet]. [cited 2020 May 3]. Available from: https://www.who.int/emergencies/diseases/novel-coronavirus-2019

4. Chen Q, Liang M, Li Y, Guo J, Fei D, Wang L, et al. Mental health care for medical staff in China during the COVID-19 outbreak. Vol. 7, The Lancet Psychiatry. Elsevier Ltd; 2020. p. e15–6.

5. Lai J, Ma S, Wang Y, Cai Z, Hu J, Wei N, et al. Factors Associated With Mental Health Outcomes Among Health Care Workers Exposed to Coronavirus Disease 2019. JAMA Netw open. 2020 Mar 2;3(3):e203976.

6. Greenberg N, Docherty M, Gnanapragasam S, Wessely S. Managing mental health challenges faced by healthcare workers during covid-19 pandemic. Vol. 368, The BMJ. BMJ Publishing Group; 2020.

7. World Health Organization. Health Workers: a Global Profile [Internet]. [cited 2020 May 11]. Available from: https://www.who.int/whr/2006/06_chap1_en.pdf

8. Wang D, Hu B, Hu C, Zhu F, Liu X, Zhang J, et al. Clinical Characteristics of 138 Hospitalized Patients with 2019 Novel Coronavirus-Infected Pneumonia in Wuhan, China. JAMA - J Am Med Assoc. 2020 Mar 17;323(11): 1061–9.

9. The Lancet. COVID-19: protecting health-care workers. Vol. 395, The Lancet. Lancet Publishing Group; 2020. p. 922.

10. Wu Z, McGoogan JM. Characteristics of and Important Lessons from the Coronavirus Disease 2019 (COVID-19) Outbreak in China: Summary of a Report of 72314 Cases from the Chinese Center for Disease Control and Prevention. JAMA - J Am Med Assoc. 2020 Apr 7;

11. MMWR. MMWR - Characteristics of Health Care Personnel with COVID-19 — United States, February 12–April 9, 2020. 2019.

12. Suwantarat N, Apisarnthanarak A. Risks to healthcare workers with emerging diseases. Curr Opin Infect Dis. 2015 Aug 25;28(4):349–61.

13. Chan-Yeung M. Severe acute respiratory syndrome (SARS) and healthcare workers. Vol. 10, International Journal of Occupational and Environmental Health. Abel Publications Services Inc.; 2004. p. 421–7.

14. Moore D, Gamage B, Bryce E, Copes R, Yassi A. Protecting health care workers from SARS and other respiratory pathogens: Organizational and individual factors that affect adherence to infection control guidelines. Vol. 33, American Journal of Infection Control. Mosby Inc.; 2005. p. 114–21.

15. Tan BH, Loh JJP, Seah SGK, Koh VWH, Lim EAS, Liaw CW, et al. Strategies adopted and lessons learnt during the severe acute respiratory syndrome crisis in Singapore. Vol. 15, Reviews in Medical Virology. 2005. p. 57–70.

16. Lau JTF, Fung KS, Wong TW, Kim JH, Wong E, Chung S, et al. SARS Transmission among Hospital Workers in Hong Kong. Emerg Infect Dis. 2004;10(2):280–6.

17. World Health Organization. Rational use of personal protective equipment for coronavirus disease (COVID-19) and considerations during severe shortages [Internet]. [cited 2020 May 3]. Available from: https://www.who.int/publications-detail/rational-use-of-personal-protective-equipment-for-coronavirus-disease-(covid-19)-and-considerations-during-severe-shortages

18. Forrester JD, Nassar AK, Maggio PM, Hawn MT. Precautions for Operating Room Team Members During the COVID-19 Pandemic. J Am Coll Surg. 2020;

19. Bhangu A, Lawani I, Ng-Kamstra JS, Wang Y, Chan A, Futaba K, et al. Global guidance for surgical care during the COVID-19 pandemic. Br J Surg. 2020 Apr 15;

20. Dashraath P, Jing Lin Jeslyn W, Mei Xian Karen L, Li Min L, Sarah L, Biswas A, et al. Coronavirus Disease 2019 (COVID-19) Pandemic and Pregnancy. Am J Obstet Gynecol. 2020 Mar;

21. Cook TM, El-Boghdadly K, McGuire B, McNarry AF, Patel A, Higgs A. Consensus guidelines for managing the airway in patients with COVID-19. Anaesthesia. 2020 Apr 1;

22. Razai MS, Doerholt K, Ladhani S, Oakeshott P. Coronavirus disease 2019 (covid-19): A guide for UK GPS. Vol. 368, The BMJ. BMJ Publishing Group; 2020.

23. Sidibé M, Campbell J. Reversing a global health workforce crisis. Vol. 93, Bulletin of the World Health Organization. World Health Organization; 2015. p. 3.

24. Liu JX, Goryakin Y, Maeda A, Bruckner T, Scheffler R. Global Health Workforce Labor Market Projections for 2030. Hum Resour Health. 2017 Feb 3;15(1).

25. Global Health Workforce Alliance, World Health Organization. A Universal Truth: No Health Without a Workforce. 2014.

26. World Health Organization. Global strategy on human resources for health: Workforce 2030.

27. Soham Bandyopadhyay, Aditi Aggarwal, Archith Kamath, Abuelgasim DKDJGBHRH, Isabel Tol, Leenah Abuelgasim, Murtaza Kadhum, Najlaa Abu Jamie, Omaima Ali, Osaid Alser, Sai Arathi Parepalli, Samuel Scott, Soph RK. Infection and mortality of health care workers worldwide from COVID-19: a scoping review protocol [Internet]. Open Science Framework. 2020 [cited 2020 Apr 17]. Available from: osf.io/jyuxb

28. Arksey H, O’Malley L. Scoping studies: Towards a methodological framework. Int J Soc Res Methodol Theory Pract. 2005 Feb;8(1):19–32.

29. Levac D, Colquhoun H, O’Brien KK. Scoping studies: Advancing the methodology. Implement Sci. 2010 Sep 20;5(1):69.

30. Micah DJ Peters, Christina Godfrey, Patricia McInerney, Zachary Munn, Andrea C. Tricco HK. Chapter 11: Scoping reviews. In: Aromataris E MZ, editor. Joanna Briggs Institute Reviewer’s Manual. 2020.

31. Tricco AC, Lillie E, Zarin W, O’Brien KK, Colquhoun H, Levac D, et al. PRISMA extension for scoping reviews (PRISMA-ScR): Checklist and explanation. Vol. 169, Annals of Internal Medicine. American College of Physicians; 2018. p. 467–73.

32. medRxiv. medRxiv.org - the preprint server for Health Sciences [Internet]. [cited 2020 May 3]. Available from: https://www.medrxiv.org/

33. Contagion Live. Coronavirus: For Health Care Workers, Risk of Infection, But Also Burnout [Internet]. [cited 2020 May 11]. Available from: https://www.contagionlive.com/news/for-health-care-workers-risk-of-infection-but-also-burnout

34. The Guardian. Healthcare workers ‘should be screened for Covid-19 every week’ [Internet]. [cited 2020 May 11]. Available from: https://www.theguardian.com/world/2020/apr/16/healthcare-workers-screened-covid-19-every-week-infectious-unethical

35. Liu Q, Luo D, Haase JE, Guo Q, Wang XQ, Liu S, et al. The experiences of health-care providers during the COVID-19 crisis in China: a qualitative study. Lancet Glob Heal. 2020;

36. Edsel B, Xu A, Salimi A, Torun N. MD Death COVID-19. medRxiv. 2020 Apr 8;2020.04.05.20054494.

37. Burrer SL, de Perio MA, Hughes MM, Kuhar DT, Luckhaupt SE, McDaniel CJ, et al. Characteristics of Health Care Personnel with COVID-19 — United States, February 12-April 9, 2020. MMWR Morb Mortal Wkly Rep. 2020 Apr 17;69(15):477–81.

38. Wang C, Pan R, Wan X, Tan Y, Xu L, McIntyre RS, et al. A longitudinal study on the mental health of general population during the COVID-19 epidemic in China. Brain Behav Immun. 2020;

